# Alternative Covid-19 mitigation measures in school classrooms: analysis using an agent-based model of SARS-CoV-2 transmission

**DOI:** 10.1101/2021.08.30.21262826

**Authors:** M. J. Woodhouse, W. P. Aspinall, R. S. J. Sparks, E. Brooks Pollock, C. Relton

## Abstract

The SARS-CoV-2 epidemic continues to have major impacts on children’s education, with schools required to implement infection control measures that have led to long periods of absence and classroom closures. We have developed an agent-based epidemiological model of SARS-CoV-2 transmission that allows us to quantify projected infection patterns within primary school classrooms, and related uncertainties; the basis of our approach is a contact model constructed using random networks, informed by structured expert judgement. The effectiveness of mitigation strategies are considered in terms of effectiveness at supressing infection outbreaks and limiting pupil absence. Covid-19 infections in schools in the UK in Autumn 2020 are re-examined and the model used for forecasting infection levels in autumn 2021, as the more infectious Delta-variant was emerging and school transmission thought likely to play a major role in an incipient new wave of the epidemic. Our results were in good agreement with available data. These findings indicate that testing-based surveillance of infections in the classroom population with isolation of positive cases is a more effective mitigation measure than bubble quarantine, both for reducing transmission in primary schools and for avoiding pupil absence, even accounting for insensitivity of self-administered tests. Bubble quarantine entails large numbers of pupils being absent from school, with only modest impact on classroom infection levels. However, maintaining reduced contact rates within the classroom can have a major beneficial impact for managing Covid-19 in school settings.

## 2. Introduction

As an increasing proportion of people become vaccinated the spread of SARS-CoV-2 becomes concentrated mainly with unvaccinated persons. Children become of particular importance in these circumstances since the benefits, to the children themselves, of vaccination are moot. While, at the time of writing, the UK government has a vaccination programme for secondary age children, primary age children have not been mass vaccinated. Factors that influence these discussions include the observation that severe COVID-19 illness is very rare in children, while there are very small risks of adverse reactions to the vaccine. Thus, the efficacy of vaccination, from the point of view of an individuals’ protection against serious disease, is equivocal. On the other hand, schools with largely unvaccinated populations may act as environments for spreading SARS-CoV-2, with the potential for development of new variants. Thus, from a public health perspective, schools are, potentially, a significant reservoir of infection. Measures to mitigate transmission can be very disruptive to learning and to the economy if large numbers of pupils and their families are required to self-isolate or have their work compromised by childcare responsibilities. In the latter context there is an urgent need to understand which policies can be pursued that reduce transmission but at the same time minimise disruption of education and collateral effects.

In this study we have developed a basic stochastic model for estimating the likelihood of the transmission of SARS-CoV-2 occurring in primary schools. There is emerging evidence that respiratory aerosols expelled by infected people are a significant mode of infection transmission [1], so indoor classrooms, which may be poorly ventilated, can represent environments with increased transmission risk. However, concentrations of virions in aerosols are substantially elevated near to an infected person, and close contacts between infected and susceptible individuals are likely to be responsible for most transmissions in many situations (see Supplementary Material Appendix A1). Our model is based on a discrete agent-based compartmental epidemiological model and involves an empirical representation of the probability of transmission due to close contacts within class-sized interactive networks. With this approach, sets of several classrooms collectively can be assessed as a way of quantifying transmission probabilities in individual schools. Statistical representation of close contacts between children and adults within school settings is derived from a study of 36 primary schools in England using teachers’ expert judgements, formally elicited in Spring/Summer 2020 [2]; in that study, close contact was defined to be a face-to-face contact within 1 metre for at least 5 minutes. It also included a comparison of anticipated contact rates in schools taking mitigation measures in the following September 2020, such as formation of bubbles, reduced class sizes and other social distancing rules, with previous, normal contact rates in ‘pre-Covid’ times.

The model results are compared with data on school attendances, self-isolation of children who are sent home due to Covid outbreaks and prevalence of Covid in both schools and communities. These comparisons enable us to check that the model produces reasonable estimations of expected infection rates and absences. However, the main purpose of our modelling approach is to compare the effects of different mitigation strategies on infection transmission rates within schools, including sending ‘*bubbles*’ or whole classes home for self-isolation periods, sending home only those pupils or adults thought to have become infected, or using testing to identify infections.

At the centre of our approach is a time-stepping agent-based epidemiological model of SARS-CoV-2 infection transmission in a classroom. The model we have developed combines a discrete compartmental epidemiological framework with a random network for daily person-to-person contacts within the classroom. In the basis case, the classroom population consists of a single teacher, a set number of pupils, and a small number of classroom teaching assistants. In relation to infection transmissions, the modelled classroom population is assumed to be isolated from other classes and persons within the school, so the present model does not include interactions with other people in the school. This is a major assumption and is best suited to primary school settings where classroom groups can be most effectively separated from one another as a part of infection risk management; this was a mitigation instituted in March 2020 for primary schools in England that were open for vulnerable children and those of critical workers and, later, as other schools in England reopened to selected age groups in June 2020. The simplifying assumption is also justified by evidence for limited mixing across different classes and year groups in UK primary schools [3].

As the model advances through time, it includes a daily chance of seeding infection within one or more of the classroom occupants from interactions with the outside community; this infection probability is a function of community incidence rate. We describe this aspect of our model in more detail below.

We then apply our model to the Autumn term (Term 1) in 2020 when UK schools first reopened to full classes. Using estimates of SARS-CoV-2 infection prevalence and the incidence rate during Term 1 2020, we illustrate the application of our model and show that it can simulate numbers of infections that are in good agreement with available data. We apply the model over this period with different mitigation measures applied and analyse the effect of these strategies on the transmission of SARS-CoV-2 within schools. We then apply our model to forecast infection in schools for Term 1 in 2021, where schools may play an important role in a third-wave of the SARS-CoV-2 epidemic, particularly in the face of the Delta variant and exclusion of children from vaccination programmes.

## 3. Methods

### Epidemiological model

We model SARS-CoV-2 transmission within a classroom of individuals using an agent-based network model, with each member of the classroom population having their own characteristics and attributes updating on interactions and over time. We adopt a compartmental epidemiological model to simplify the states of agents, and to accommodate transmission from pre-symptomatic and asymptomatic individuals, the progression of infection is described using a variation of a stochastic compartmental epidemiological model, which includes explicit time variation in the transmissibility of SARS-CoV-2 based on time since infection. Specifically, we use a compartmental model with the following compartments: Susceptible (*S*), Exposed (*E*, which includes infectious individuals), Unwell (*U*, where an individual has symptoms), Quarantined (*Q*), and Recovered (*R*). We adopt a discrete-time stochastic modelling framework, with updates to the compartmental model occurring each day. Further details of the agent-based model are given in Supplementary Material Appendix A2.

The Susceptible population are those who have not become infected with the SARS-CoV-2 virus. On becoming infected, an individual moves to the Exposed compartment and begins a period of incubation during which there is no external indication of infection, but where there is a possibility of infection transmission in contacts. For symptomatic individuals the incubation period ends with the individual experiencing symptoms and moving into the Unwell compartment. At this stage we assume that symptoms of infection would be recognized by the infected individual or observed by others and, if such symptoms are detected or confirmed, usually by testing, then an Unwell individual will be Quarantined. Individuals remain quarantined for a minimum quarantine period, and for as long as they are unwell. However, others may be asymptomatic or have symptoms so mild as to be unobserved, so these persons are not migrated to the Unwell category in the model. The Recovered compartment collects infected individuals following their period of being Unwell, or for the duration of the infectious period for asymptomatic cases, at the which stage they are assumed to have acquired immunity from further SARS-CoV-2 infection for the simulation period.

Using evidence derived from large datasets on the timing of secondary infections with the onset of symptoms of primary cases [4], we model infectivity through time-dependent transmission probability of an index case with a contact. The transmission probability increases during the incubation period from zero at the time of infection, peaks slightly before the time of symptom onset (for symptomatic cases), and subsequently decreases over time [4]. In [4] it is inferred from data that there is a distribution of the duration of incubation period of SARS-CoV-2 and that the duration of pre-symptomatic infectious period is related to the duration of the incubation period, with individuals who have long incubation periods tending to have an earlier and longer lasting pre-symptomatic infectious period. Future variants of the virus could alter these distributions.

These time-dependencies are encoded in an expression for the relative probability of transmission in a contact within the class that is a function of time (*t*, in days) since symptom onset [4], denoted here by *p*_*r*_(*t*|*t*_*i*_), which depends on the incubation period of the infected individual (*t*_*i*_, in days). For the distribution of the incubation period we use a log-Normal distribution, *t*_*i*_∼log*N*(1.63, 0.5) [5]. For the relative probability of transmission, we adopt the parametric model determined from data by [4],

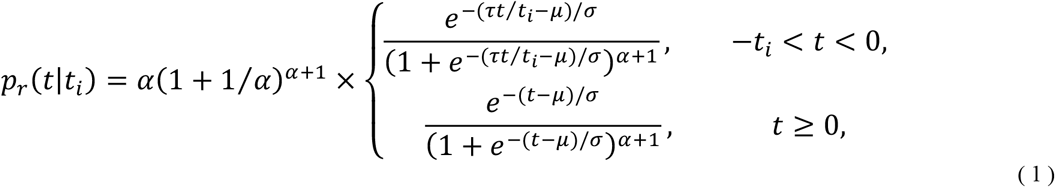

where the parameters *μ* = −4 days, *σ* = 1.85 days, *α* = 5.85 are best-fit parameters as estimated by [3], and *τ* = 5.42 days is the mean incubation period [5]. Note, this function is not a probability density function, rather it is applied as a time-varying weighting to the transmission probability in a contact, and the pre-factor (*α*(1 + 1/*α*)^*α*+1^) is specified so that 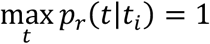.

The time variation of the relative transmission probability is illustrated in Figure 1 for an unusually short (*t*_*i*_ = 2, *P*(*t*_*i*_ < 2) < 0.05), a typical (*t*_*i*_ = 5, *P*(*t*_*i*_ < 5) = 0.48) and an unusually long (*t*_*i*_=12,*P*(*t*_*i*_ > 12) < 0.05) incubation period.

**Figure 1.**
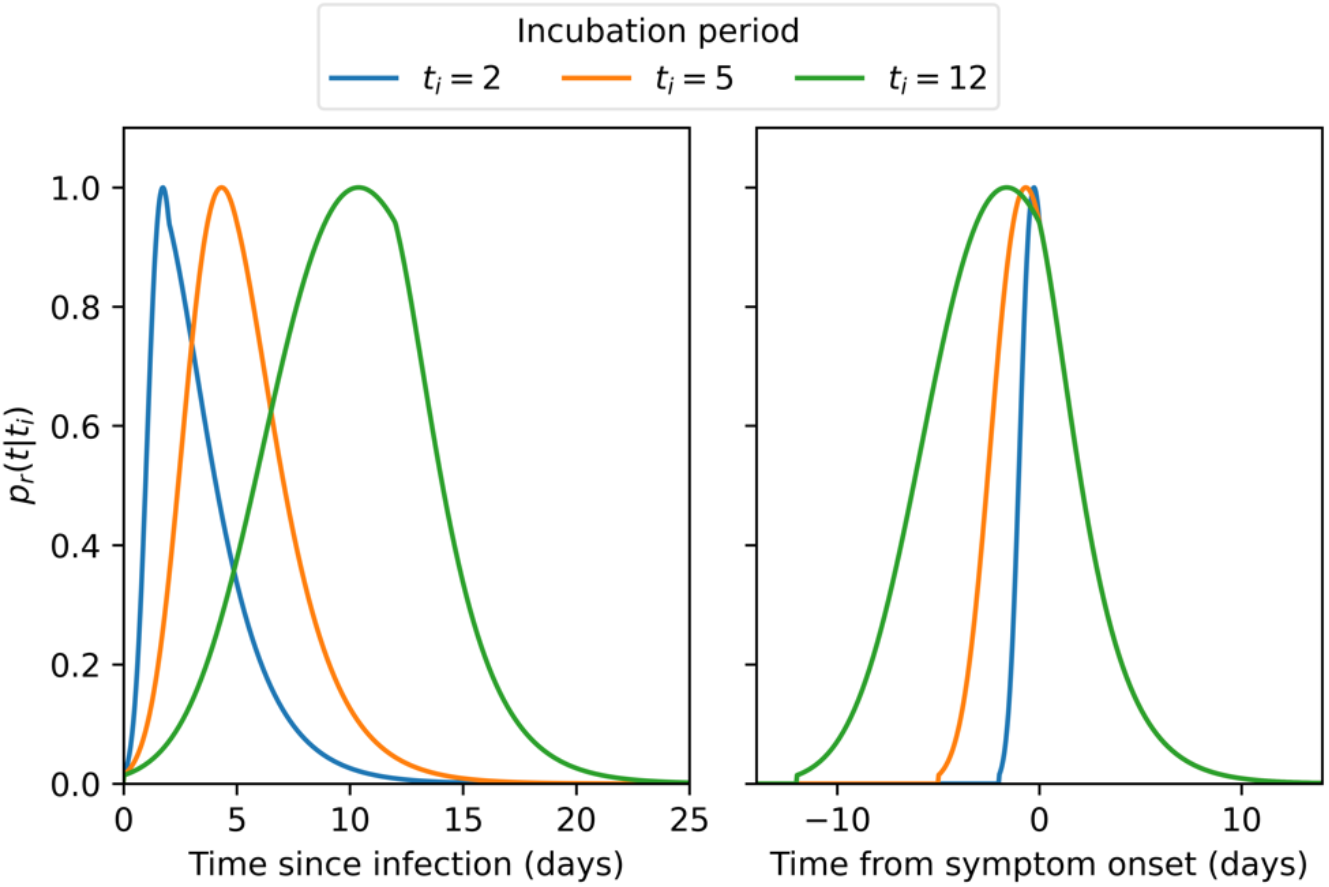
Time variation of the relative probability of transmission from an infected individual with incubation period t_i_. Probability of transmission increases from the time of infection, peaks close to the time of onset of symptoms, and then decreases over the duration of infectivity. Individuals with long incubation periods have longer pre-symptomatic infection periods, but following symptom onset the decrease in transmission probability is independent of t_i_.

The absolute probability of transmission in a contact is found by scaling the relative probability of transmission *p*_*i*_ (*t*|*t*_*i*_) by a fixed peak transmission probability, denote by *p*_*max*_, which differs for adults and children [6-8], for symptomatic or asymptomatic cases [9], reflects mitigation measures, and is altered to model differences in transmissibility for SARS-CoV-2 variants.

The classroom population can comprise both symptomatic and asymptomatic individuals, distinguished as these who will (respectively, will not) develop identifiable symptoms if they are infected with SARS-CoV-2. The specification of symptomatic and asymptomatic individuals occurs on initialization of the model by random Bernoulli draws with a prescribed probability. For asymptomatic cases, we adopt the same time-varying relative probability of transmission by specifying a notional incubation time (drawn from the log-Normal distribution), but the absolute probability of transmission in a contact is reduced relative to symptomatic cases by 30% (Table 1).

**Table 1.**
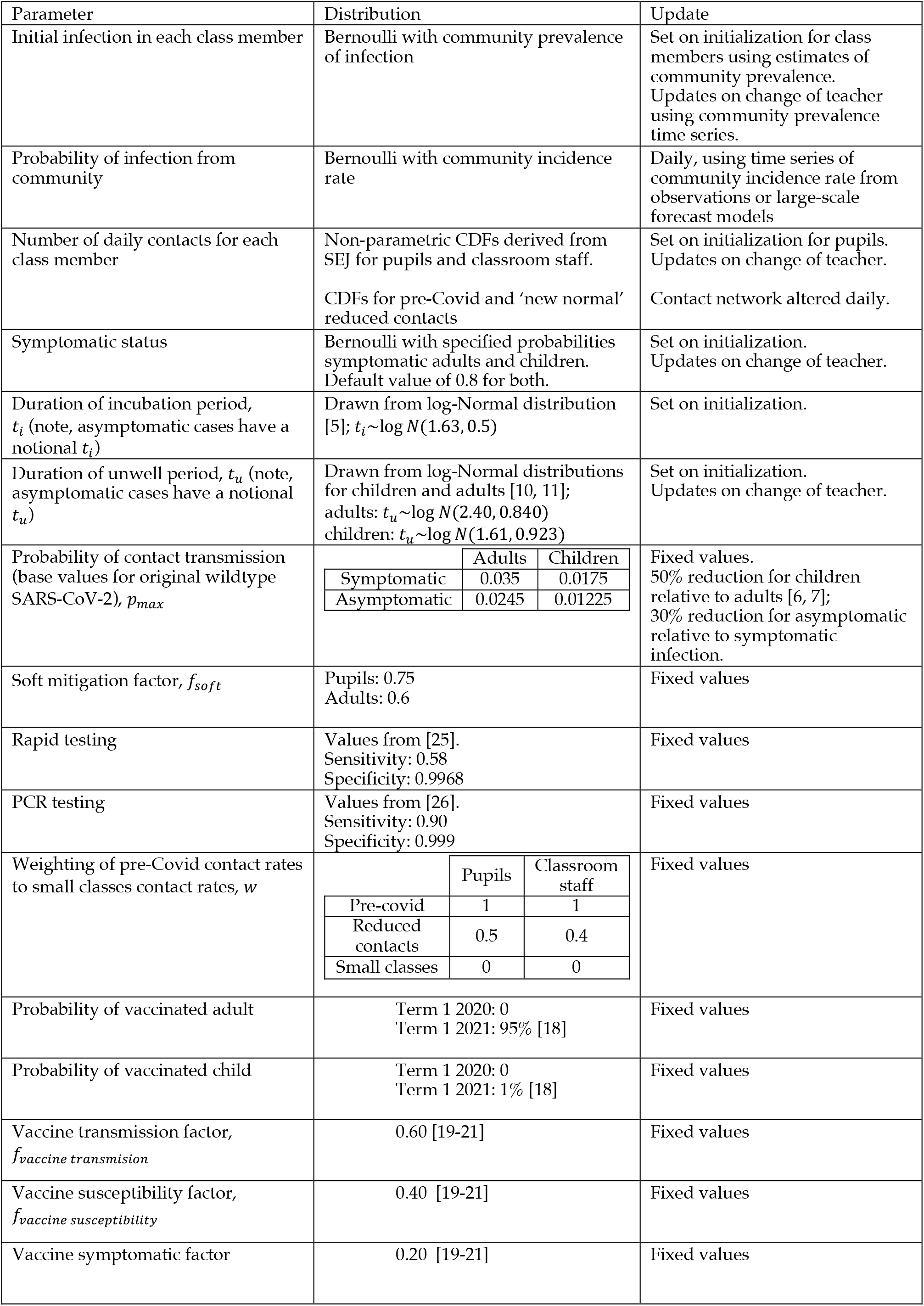
Stochastic and fixed parameters of the model.

Symptomatic individuals become unwell following the incubation period. The duration of symptoms varies for adults and children [e.g., 10, 11] with adults typically having longer lasting symptoms (median duration of 11 days) compared to children (median duration of 5 days for children aged 5— 11 years [11]). However, there is pronounced variation around the median duration which we model using a log-Normal distribution. This is a two-parameter distribution, specified by using the reported proportion of people having symptoms extending beyond 28 days in [10, 11] (13.3% for adults and 3.1% of primary school aged children).

An additional ingredient of the population dynamics is that quarantined teachers must be replaced by a substitute teacher, drawn from outside the initial class group.

Initialization of the model places the classroom members into the *SEUQR* compartments. A Bernoulli trial is performed for each individual using a community infection prevalence, specified from observation or national scale model-based projections, with ‘*failure*’ resulting in the individual placed into the Susceptible compartment. A ‘*success*’ in the Bernoulli trial corresponds to an individual with infection on initialization, so random draw is made to determine their elapsed time of infection and they are placed into the appropriate E, U or Q compartments on this basis. We note that initialization may result in classrooms without an infected individual, particularly when community prevalence is low. Additionally, we can model pre-existing immunity, either through previous infection or vaccination, with Bernoulli trials that adopt the probability of an individual being vaccinated or previously infected.

### Transmission into the classroom

In our model there are two pathways for susceptible members of the classroom to become exposed: contact with an infectious member of the classroom, or through interactions outside the classroom (referred to here as ‘*community transmission*’). To model the infection from an out-of-classroom interaction, we begin each daily time step with a Bernoulli trial for each susceptible classroom member. The probability of any susceptible classroom individual being exposed to community transmission is specified using an estimate of the daily incidence rate of new infections at the appropriate time. This approach overestimates community transmission as the individual is in school for much of the day, and classroom transmissions will contribute to the community incidence rate estimates. However, without detailed estimates of incidence rates in different settings, it is difficult to deconvolve these effects, and our approach is a pragmatic comprise, recognizing that time in school represents approximately 26% of waking time in any school week, and our model does not explicitly include effects that might promote incidence of infection connected to schools (such as increased social mixing at the start and end of the school day).

### Classroom transmission network

Transmission of infection within the classroom requires a more sophisticated model. Potential pathways for SARS-CoV-2 transmission include physical contact, infection by virus contained in large respiratory droplets, contagiously from contaminated surfaces (fomites), and through inhalation of infectious aerosols (airborne transmission) that could act over long distances [12]. For all of these routes, close contacts are likely to substantially increase the probably of infection transmission (see Supplementary Materials Appendix A1 for a discussion of airborne transmission). We therefore base our transmission model on close contacts between individuals, using a random network model of contacts within a classroom. The number of contacts for each individual is specified from stochastic draws from the distribution of daily classroom contacts [2]. Thus, each class member has their own number of daily contacts with others in the classroom.

It is known that contact patterns in classrooms can be highly variable, affected by both individual and classroom behaviours [2]. Additionally, behavioural mitigation measures were instituted in March 2020 to reduce contacts within schools. To model the number of contacts, we use the results of a Structured Expert Judgement (SEJ) elicitation study [2] conducted in spring/summer 2020 in the UK. SEJ uses the collective knowledge of experts (here school teachers) to estimate quantities of interest and their uncertainties, and can be performed rapidly during crises to provide essential information to models and policy makers [13]. Experts’ responses to a set of test questions with knowable results are used to score experts in terms of their individual statistical performance and their target item judgements are then combined jointly, using these performance scores, to derive a pooled ‘Decision Maker’ [13]; the latter represents associated group-wise judgement uncertainties through empirical cumulative distribution functions (eCDFs) [14].

In our model, empirical cumulative distribution functions for the number of contacts in classrooms are those derived from pooled expert responses in an elicitation for primary school pupils and classroom staff [2]. That study determined different contact rates in: (a) the pre-Covid classroom, and then (b) following implementation of bubbles and other measures to reduce the number of contacts [2]. The corresponding eCDFs are illustrated in Figure 2a, showing substantial reduction in the number of contacts once mitigation measures are imposed. The mean number of contacts for pupils is reduced from 16.7 per day (std. dev. 16) in pre-Covid classrooms, to 6.4 per day (std. dev. 5.4) when there are small nunbers in the class. We note that the mean daily pupils’ contact rate is similar to the value estimated by the POLYMOD social contact survey [15] (mean of 18.2 contacts per day, std. dev. 12.2 for children aged 10—14 years).

**Figure 2.**
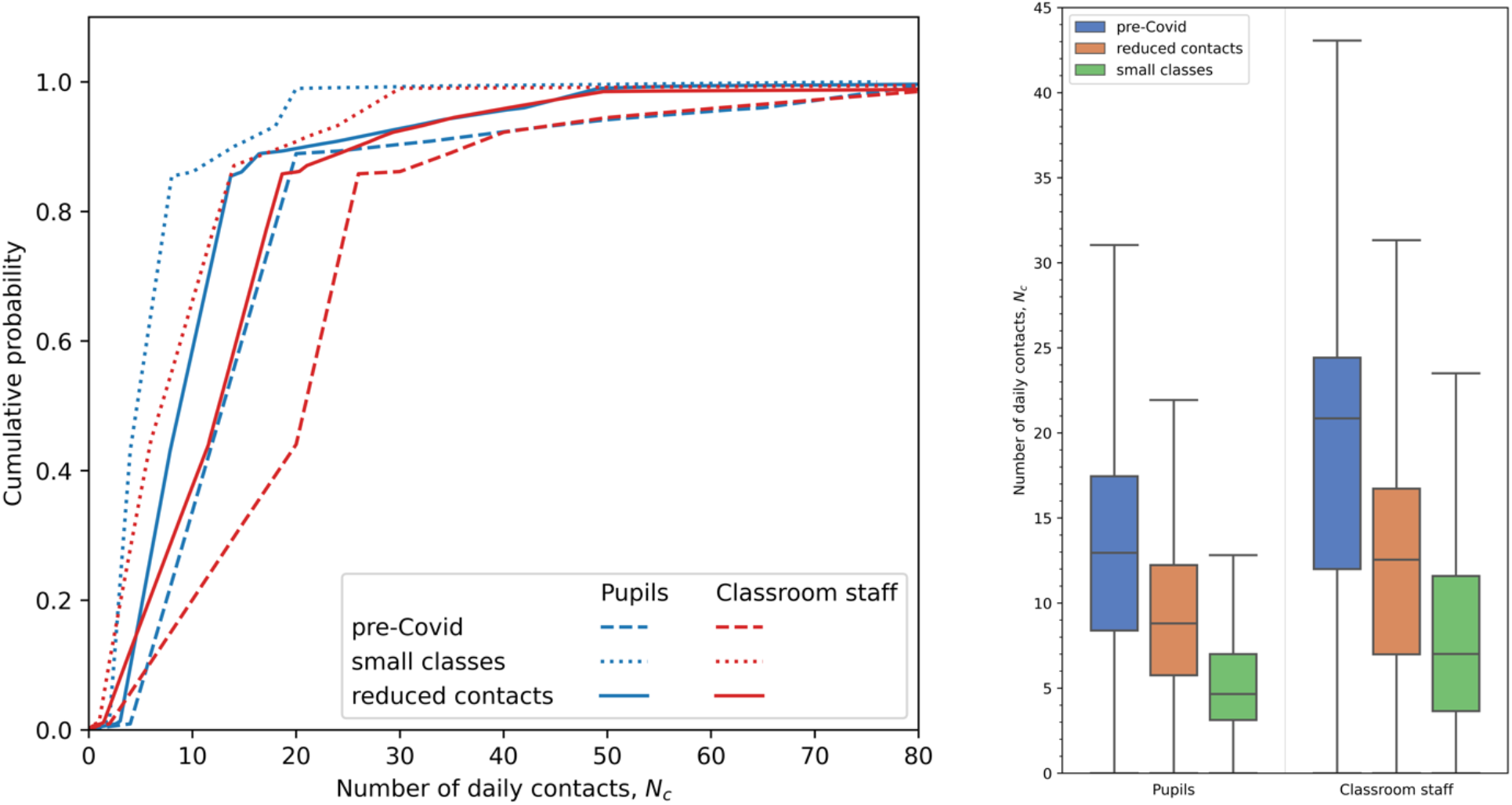
a) Empirical cumulative distribution functions (eCDFs) for the number of daily contacts in primary school classrooms. The eCDF derived from Structured Expert Judgement for pupils (blue) and classroom staff (teachers and teaching assistants; red) are shown for pre-Covid times (dashed lines) and following opening of schools to small numbers of children in June—July 2020 (dotted lines). The cumulative distribution function constructed from a weighted combination of the pre-Covid and small classes eCDFs are shown (solid lines) b) Box and whisker plots (with outliers removed) comparing the number of daily contacts for pupils and classroom staff in pre-Covid times, for small classes, and modelled for full classes under conditions to reduce contacts.

The contact data estimated by SEJ in [2] were obtained under the unique circumstances of June and July 2020 when between 30% and 40% of children had returned to school and the class sizes were greatly reduced, making social distancing measures much easier to implement. However, with full return to school we would not expect the same reduction in contact rates. In study [2] teachers were asked to estimate whether the contact numbers would be affected. Their collective view (Table 7 in reference [2]) is that the contact numbers would be between their estimates in normal pre-Covid times and in new normal times.

To model the reduction in the number of contacts in fully-opened schools, we construct a cumulative distribution function (denoted by *P*_*r*_ (*N* = *n*)) as a weighted linear combination of the eCDF in pre-Covid classrooms in normal times (denoted by *P*_*n*_(*N* = *n*)) and the eCDF in the small classrooms of June—July 2020 (denoted by *P*_*s*_(*N* = *n*)), with *P*_*r*_(*n*) = *wP*_*n*_(*n*) + (1 − *w*)*P*_*s*_(*n*) where *w* is the weighting of the pre-Covid contact distribution. Contact rate distributions are constructed in this way for both pupils and teachers, with differing weights. In [2] teachers expected that daily pupil contacts would be half-way between the small classroom and normal pre-Covid values when classrooms become more highly occupied, and we therefore take a weight *w* = 0.5. However, teachers considered that their own numbers of daily contacts could be more strongly reduced in the fully open schools, so we take *w* = 0.4 to place greater weight on the small classroom distribution. With these weightings, the mean number of contacts per day is 11.6 (std. dev. 10.9) for pupils and 15.1 (std. dev. 15.9) for treachers. These summary values are similar to the contact rates for children estimated using surveys over Term 1 (September—December) of 2020 when schools fully reopened (mean number of 15.1 contacts per day) [16], albeit with different definitions of a contact (in [2] face-to-face contact within 1 m for 5 min or more is used, whereas [16] define direct contact as an interaction where “*least a few words were exchanged or physical contact was made*”).

At the start of each school day in the modelled sequence, we build a stochastic network of contacts for each class member who is not quarantined. The network is constructed using the ‘*Configuration Model*’ approach [e.g. 17], allowing multiple edges but removing self-loops through a sequence of random ‘*rewirings’* (i.e. self-loop edges are randomly switched until a valid configuration is achieved). Preserving multiple edges allows us to model repeated contacts between individuals. The Configuration Model produces a random graph with a specified degree distribution in which nodes of the network are linked randomly using specified numbers of edges. This means that gregarious individuals with high numbers of contacts are more likely to be connected to other highly contacting individuals. However, the contact network is rebuilt each day, to reflect changing contact patterns and the removal of class members into quarantine. The Configuration Model requires an even number of connections; in cases where an odd number of edges is generated by the stochastic initialization of the degree distribution or from removal of nodes to quarantine, then the degree of a randomly selected node is increased by one. On non-school days, there are no classroom contacts, but community transmission can occur, and the agents continue to progress through the *SEUQR* compartments.

We note that in realty contacts in a class are unlikely to be completely random. Children for example form friendship groups which may mean that poorly connected networks may exist. While the Configuration Model with degrees specified by the contact rates distributions results in small groups with large numbers of interactions, the networks are typically connected. The Configuration Model does not exclude disconnected sub-graphs, but we find they are rare occurrences in our simulations. Addressing the detailed structure of classroom contact networks would make an interesting development of the model.

Figure 3 shows examples of the contact networks, illustrated using the ‘adjacency matrix’ (i.e. the number of edges linking each pair of nodes, corresponding to the number of daily contacts between two individuals in the classroom). An example of a randomly generated network is shown for both contacts rates drawn from the pre-Covid and reduced contacts distributions. For pre-Covid contacts (Figure 3a) the most gregarious member of the class has 85 daily contacts, and there are four individuals with more than 50 contacts per day (pupils numbered 0, 6, 21 and 30). These highly contacting individuals have many interactions within this subgroup, with e.g. pupils 21 and 30 having 11 contacts on this day. When reduced contacts rates are employed (Figure 3b) the most contacts decreases to 48, and only a single individual has more than 23 contacts in the day, and the most contacts between pairs of individuals, between pupils numbered 22 and 28, occurs only 4 times on this day. There are no disconnected sub-graphs in either of the examples shown.

**Figure 3.**
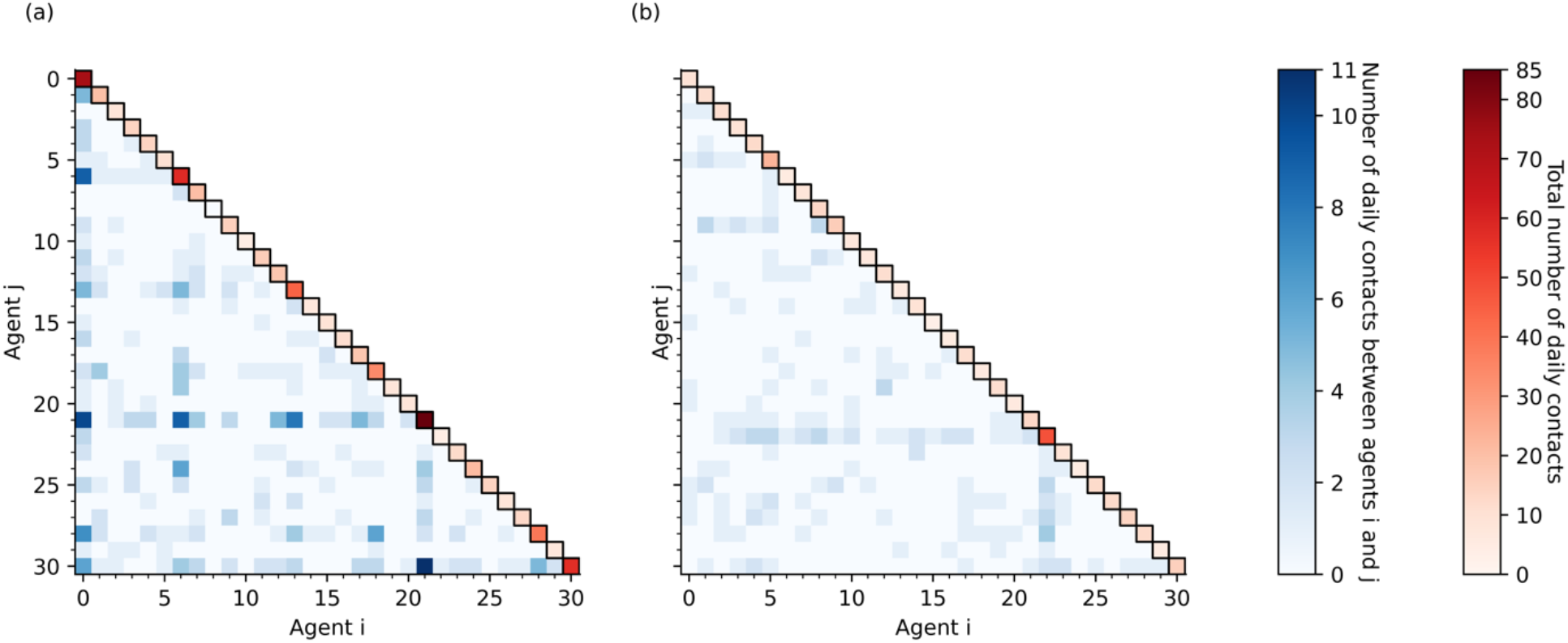
Examples of adjacency matrices (illustrated with blue shades) for stochastic contact networks produced using the Configuration Model with the total number of contacts for each agent (the degrees of each node, shown in red shades on the leading diagonal) drawn from (a) pre-Covid contact rate distributions and (b) the reduced contacts rate distributions. The thirty ‘agents’ in the classroom are numbered 0—30, with agent 30 corresponding to the teacher. Note the same colour scales are applied for both pre-Covid and reduced contacts cases.

If contact occurs between an infectious individual with another who is susceptible, there is the possibility of transmission. This is modelled as a random Bernoulli trial, with differing probabilities of transmission depending on whether the infectious individual is symptomatic (higher transmission probability) or asymptomatic (lower transmission probability), and with transmission probability varying in time. In a contact between a Susceptible person in the classroom with an infectious person, we determine the infection transmission probability as

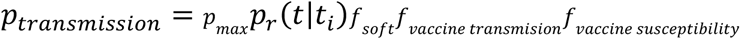

where *f*_*soft*_*f*_*vaccine transmision*_*f*and *f*_*vaccine susceptibility*_ are factors (discussed further below) that reduce transmission probability due to application of ‘soft’ mitigation measures. and effect of vaccination of infectivity and susceptibility. Note, this approach combines the infectiousness of the index case with the susceptibility of their contact into a single probability of transmission. This is a simplifying assumption in the model, but includes individual susceptibility to SARS-CoV-2 for the population in the model. The SEUQR compartments are updated at the end of daily timesteps, so transmission can occur before recovery.

An additional requirement is that the classroom must have a teacher present. If the teacher is quarantined due to illness or a positive test result, a temporary replacement is added to the classroom population. The substitute teacher has their own number of daily contacts, drawn from the SEJ distribution, and their stage in the *SEUQR* progression is determined by Bernoulli trial with the infection prevalence at the time of replacement, noting that a valid substitute teacher cannot be showing symptoms (i.e., Unwell) nor in Quarantine, but could bring infection into the classroom population. If the permanent teacher is released from Quarantine, or their time of illness elapses (i.e., they progress to Recovered), then they are returned to the classroom and the substitute teacher is removed. Should the temporary teacher become Unwell or Quarantined, they are replaced by a different, substitute teacher.

Examples of three random contact networks produced using the Configuration Model are illustrated in Figure 4 for a small classroom of ten pupils (enumerated from 1 to 10) and a single teacher (labelled as ‘T’), simulated over three days. The degree distribution is fixed; pupils 1 and 2 are the most gregarious with five contacts each day, followed by pupil 5 with five contacts on days 1 and 3 but four contacts on day 2. There are duplicated contacts (i.e., multiple edges) between pupils in each network, which present two opportunities for transmission between persons if one of the individuals is infectious and the other susceptible (which occurs in Figure 4c between pupils 1 and 2).

**Figure 4.**
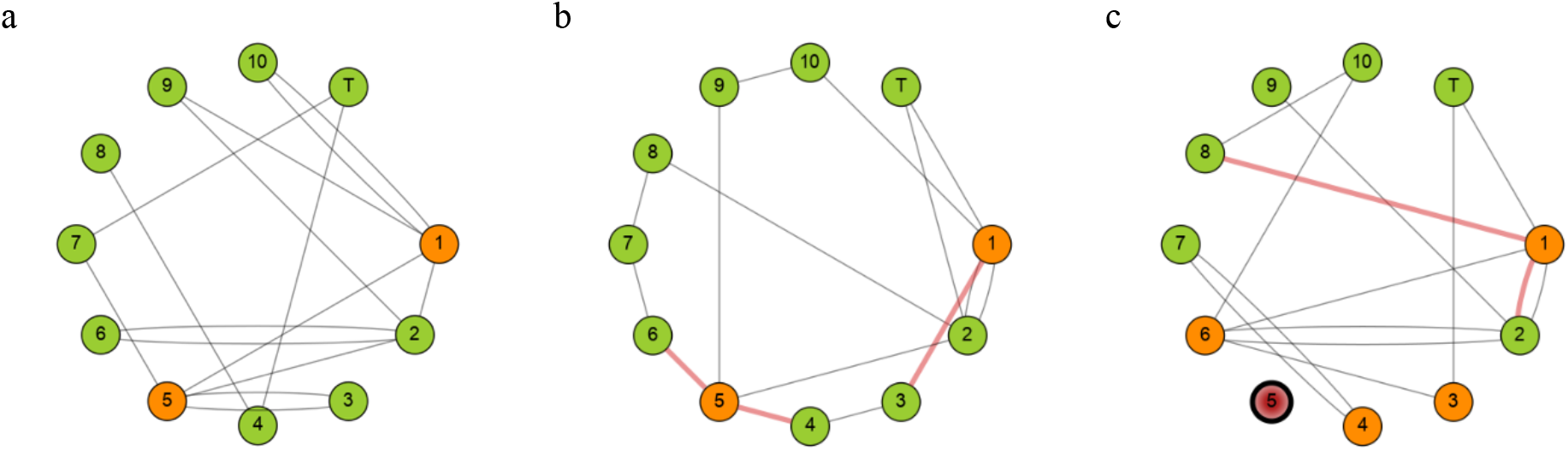
An example of infection transmission within a random updating contact network in a small classroom of ten pupils and one teacher. The degree distribution is fixed, and the edges are determined using the Configuration Model with self-loops removed by random rewiring. Multiple edges are allowed, as illustrated in a where e.g. there are two edges between nodes 1 and 10. The panels represent a progression over three days. Green nodes indicate Susceptible class members, orange nodes indicate Exposed members, and red nodes indicate Unwell individuals. Nodes that are boldly edged (node 5 in c) are quarantined. Red coloured edges indicate infection transmission between infectious and susceptible members. Note the parameters for infection transmission in this example are not representative of SARS-CoV-2.

Figure 4 also illustrates the transmission of infection across the network, with a daily progression from panels a to b to c. Note that the parameters related to infection transmission in the example in Figure 4 are taken to be much larger than reasonable to model SARS-CoV-2 and have been chosen simply to illustrate the way the model updates. On the first day (Figure 4a), there are two exposed individuals (pupils 1 and 5), and their contacts represent potential pathways for infection, with actual transmission determined by Bernoulli trials. In this example, the random trials result in no transmissions. However, on the second day (Figure 4b) the two exposed pupils do transmit the infection across some of their contacts; of the five contacts with pupil 1, there is one transmission to pupil 3, whereas two contacts of pupil 5 result in transmission to pupils 4 and 6. Therefore, on the third day, pupils 3, 4 and 6 are progressed from the Susceptible into the Exposed compartments. Further infection transmission occurs on day 3, with two contacts of pupil 1, but, by chance and there is no infection transmission in contacts of the other exposed individuals. Additionally, on day 3, the pupil 5 has developed symptoms and is progressed into the Unwell compartment. In this example, the Unwell individual is Quarantined away from the classroom and has no connections with other members of the classroom.

### Vaccination uptake and effectiveness

As vaccination programmes continue, there is a need to include the effects of vaccine on SARS-CoV-2 infection, transmission and symptoms. In the UK, vaccination began in October 2020 with high-risk groups and has proceeded downwards through age-groups. Therefore, in modelling school infection in Term 1 2020, there are very low levels of vaccination for pupils and teachers. However, vaccine uptake has been high, so that by the end of the 2020—2021 school year 80% of working age people are estimated to have received two vaccine doses [18]. Primary aged children are unlikely to have received vaccine unless they have additional risk factors. To include vaccination in our model, on initialization of the class members, we perform Bernoulli trials using estimates of the vaccination uptake proportion.

We model three effects of the vaccine: (i) a reduction of the probability of a vaccinated individual becoming infected; (ii) a reduction of the probability of an infected vaccinated individual transmitting the infection in a contact; (iii) a reduction of the probability of an infected vaccinated individual developing symptoms. The different available vaccines have different effectiveness in each of these [19, 20]. Here we use fixed values for scaling factors applied for the reduction in probability of infection, reduction in probability of transmission, and reduction in probability of symptoms for vaccinated members of the population. Therefore, for susceptible vaccinated individuals, their probability of becoming infected by both community transmission and classroom contact is reduced by a ‘*Vaccine Susceptibility Factor*’. For an infected vaccinated individual, their probability of onward transmission in a contact is reduced by a ‘*Vaccine Transmission Factor*’, and their probability of developing symptoms is reduced by a ‘*Vaccine Symptoms Factor*’. These are assigned plausible values based on [19-21] (Table 1).

### Mitigations

To understand the effectiveness of mitigation measures in reducing SARS-CoV-2 transmission and infection prevalence within the classroom population, we consider several ‘*hard*’ and ‘*soft*’ approaches. Soft measures include enhanced cleaning and hygiene, as well as the reduction of mixing of separate classrooms (e.g., through staggered entry, breaks and lunch times; multiple and separated entry points to the school, etc.). We do not model these explicitly as the efficacy of some of these measures is limited [22] and is not well known and would substantially increase the complexity of the model and its interpretation. Instead, we apply soft mitigation factors that reduce the absolute probability of transmission in a contact by a fixed factor and specify different values for adults and children.

Hard mitigations correspond to isolation of classroom members away from the school and result either from the identification of symptoms or from testing surveillance to detect pre-symptomatic infection or their contacts. We have discussed above the removal of symptomatic individuals from the classroom as they reach the Unwell stage of the *SEUQR* progression. In our model, we also allow for two other hard mitigations: ‘*bubble quarantine*’ of the entire class, and ‘*regular rapid testing*’ of all individuals.

The ‘*bubble quarantine*’ approach is severe, with the entire class placed into Quarantine for a specified time period if there is a confirmed infection case in the class (e.g., if a classroom member becomes unwell). During this time, there are no classroom contacts. However, classroom members may still acquire infection through community transmission and continue to progress through the S, E, U, R compartments. For Susceptible members in quarantine, the probability of becoming infected through community transmission is likely to be reduced as their contacts will be substantially reduced. However, the as infection transmission within households is the primary pathway for infection [23], the reduction in community transmission is likely to be small, and for simplicity we do not include in our model as it would introduce a further highly uncertain parameter. Any individual who becomes Unwell during Quarantine restarts their quarantine period. On completion of Quarantine all classroom members (except those who are Unwell) are returned to the classroom. A individual in Quarantine can become Unwell if they are infected, and then remain in Quarantine until they move to the Recovered compartment and return to the classroom. Note, there may be asymptomatic or pre-symptomatic infected individuals returned to the classroom following bubble quarantine.

An alternative and less severe approach is ‘*regular rapid testing*’. This surveillance method adopts the regular lateral flow testing of all the classroom population on specified days. The rapid testing is capable of identifying asymptomatic and pre-symptomatic infection, allowing early quarantine by advancing members from the Exposed compartment to Quarantine. However, all tests have the potential to return misleading results, which may be particularly acute when administered by untrained people [24]. Therefore, we model the application of rapid testing using a stochastic approach. On each test day, each classroom member undergoes a Bernoulli trial: for Exposed individuals who have the infection, the trial adopts the rapid test sensitivity (the true positive rate, i.e., the test probability of correctly detecting infection in an infected subject); for Susceptible individuals who are not infected, the trail adopts the rapid test false positive rate (i.e., the test probability of incorrectly detecting infection in an uninfected subject = 1 − specificity). A ‘*success*’ in each trial corresponds to a positive test result and leads to the individual moving into Quarantine. However, for uninfected individuals this is an incorrect test result, and the Susceptible member is unnecessarily removed from the classroom.

The sensitivity of rapid testing in the UK can be high if administered correctly [24]. However, in studies of lateral flow tests self-administered by untrained members of the public, the sensitivity can be as low as 0.58 [25]. We adopt this value here. We have not found similar values for the test specificity for self-administered lateral flow tests, so adopt the reported value of 0.9968 despite our expectation that this value is inflated.

We consider a further approach to reduce the number of classroom members unnecessarily quarantined by false positive rapid tests through the use of confirmatory PCR testing. In our model system we only include PCR tests as confirmatory to a lateral flow test. The PCR tests are more accurate (high sensitivity and high specificity) and can detect infection sooner after transmission, but require laboratory analysis, so are not well suited for regular testing of large populations. Here we allow for them to be used to confirm (or overturn) a positive rapid test result. The PCR test results do not appear immediately; rather, there is a lag while the test is processed, during which time the test-positive subject remains in Quarantine. The sensitivity and specificity of the PCR tests are reported to be much higher than rapid lateral flow tests [26].

### Model parameters and stochastic ensemble

The parameters in the model are presented in Table 1. Many of them are uncertain; where possible we have adopted published values as noted in Table 1 (although many studies have not completed peer-review at the time of writing), and otherwise we have taken reasonable values. We have not attempted to ‘fit’ parameters to data, but such a study would be valuable. Therefore, our reported values are best used as comparison between the different scenarios we simulate rather than detailed hind-or forecasts of absolute numbers of infections, although, as we show below, our results are broadly comparable to observed infection levels.

There are several stochastic components in the model, which we also summarise in Table 1, and therefore we must perform stochastic ensemble simulations of sufficient size to explore the space of possible trajectories of the infection dynamics within a classroom. In this study, we take ensembles with 100,000 realizations of isolated classrooms. Note, in school year 2020—2021 there are 156,843 state primary school classrooms and 4.18 million pupils in England [27] (see also [28]). Ensembles of 100,000 samples of our hypothetical class of 30 pupils provide indicative traits and trends in distributional variability; if we invoke the ergodicity assumption, while admitting national schools classes, *en masse*, will have a variety of pupil numbers and will have other factors in play (inner city -v-rural; economic differences, etc), then our 30-pupil class infection distributional profile can be adopted as first order representative of all real classes *faute de mieux*. On this basis, we expect that findings from the realizations will scale to transmission at a national level.

## 4. Retrospective analysis

The beginning of the 2020—2021 school year in September was the first time that there was ‘full’ attendance in schools since the start of the SARS-CoV-2 epidemic in the UK. The prevalence of infection in the UK in September 2020 was relatively low after the aggressive mitigation measures put in place in spring and early summer, but relaxation of restrictions over the traditional school summer holiday period in August resulted in increasing incidence of infection across the UK as schools reopened in September, a trend that persisted through Term 1 (Tuesday 1 September to Friday 23 October 2020) as shown in Figure 5. Data from the Office for National Statistics [29] gives infection prevalence of 0.09% in England for all ages, and 0.11% for children (age 2 to school year 6) on 1 September 2020, increasing by about an order of magnitude, to 1.10% for all ages and 0.87% for children, by 23 October 2020. The incidence rate (per 10,000 population per day) was 0.74 on 1 September, peaked at 8.44 on 21 October 2020, falling slightly to 8.41 on 23 October 2020. At this time, vaccine uptake was low (the national vaccination programme had not started in September and was targeted at high-risk groups through October 2020) so we do not include vaccination in our simulations.

**Figure 5.**
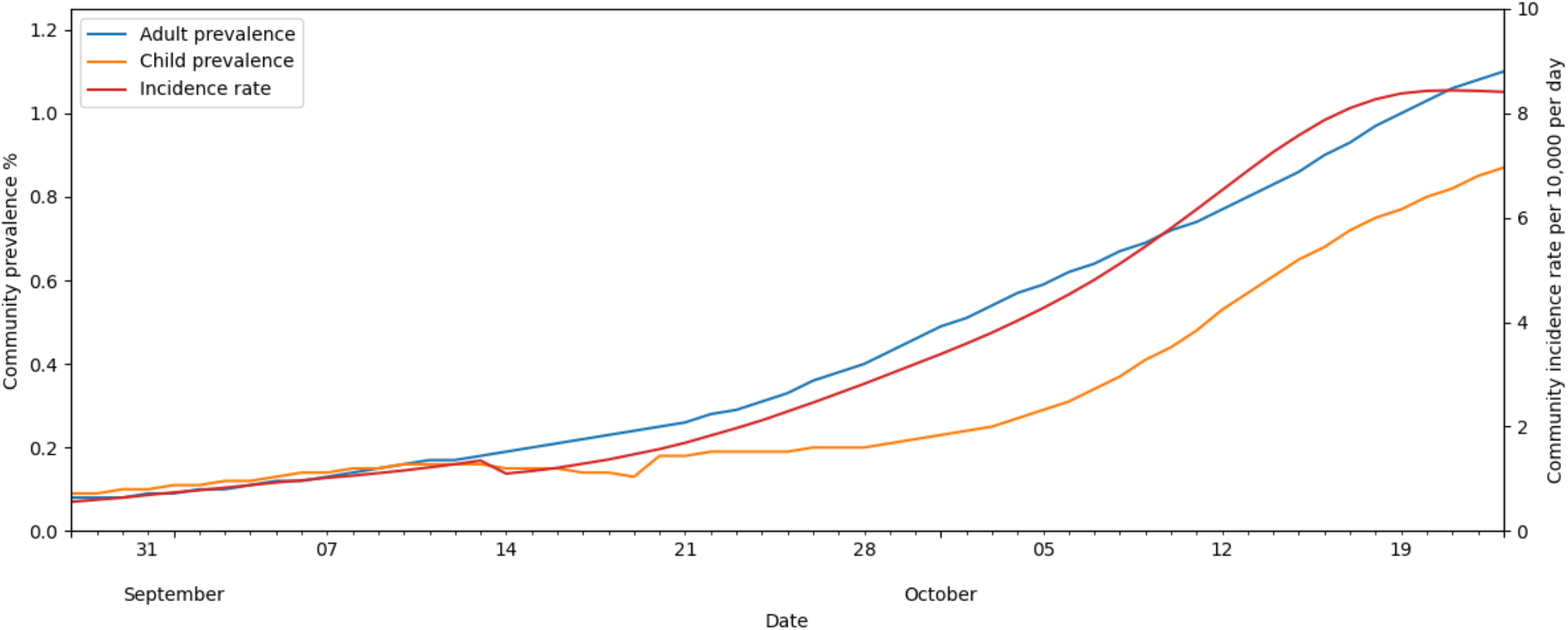
Time series of Covid-19 infection prevalence for adults (blue line) and children (age 2 to school year 6; orange line) and the community incidence rate (red line) over Term 1 (Tuesday 1 September to Friday 23 October 2020). Data from Office for National Statistics [29].

### School collections

Although we model SARS-CoV-2 in a single isolated classroom, the level of infection in larger collections of classrooms (e.g., classes in a school, and schools together in a local authority area) are often the most pertinent issues for decision makers. We therefore model a school as a collection of classrooms, and several schools collected together, through sampling with replacement from a stochastic ensemble of simulations of independent individual classes. Specifically, we generate an ensemble of simulations and randomly select realizations from this ensemble to produce a collection of classrooms to represent a single school, and a collection of such schools to model e.g., a local authority area or academy network. An ensemble of such classroom collections is readily derived by resampling with replacement from the single-classroom ensemble so that statistically meaningful metrics can be derived. We take this approach when comparing mitigation strategies.

We model a ‘typical’ primary school classroom, consisting of 30 pupils and a teacher, over Term 1 in an ensemble with 100,000 realizations. At this time there were no ‘hard’ mitigation measures, and pupils were only asked to self-isolate if they (or a close contact) developed symptoms. However, soft mitigations including enhanced cleaning and reduction of classroom contacts were in place. We therefore do not incorporate hard mitigation measures in the model simulations but build our contact networks using the reduced contacts eCDFs from SEJ, while presuming pupils are quarantined if they become unwell.

In Figure 6 we illustrate the dynamics of classroom transmission over Term 1 by plotting the timeseries of the number of infected pupils in a classroom for each of the ensemble realizations. The ensemble members have been ordered by the maximum number of infected pupils, which peaks at 14 across all ensemble members. A slight majority of realizations (54%) have no infected pupils across the whole of Term 1. The numbers of infected pupils increase towards the end of Term 1, following the trend in the community incidence rate and, we infer, reflects seeding of infection in the classroom largely from outside community transmission. Figure 7 shows the number of infections within the ensemble that results from either community (Figure 7a) or classroom transmission (Figure 7b), and their correlation with the total number of infections.

**Figure 6.**
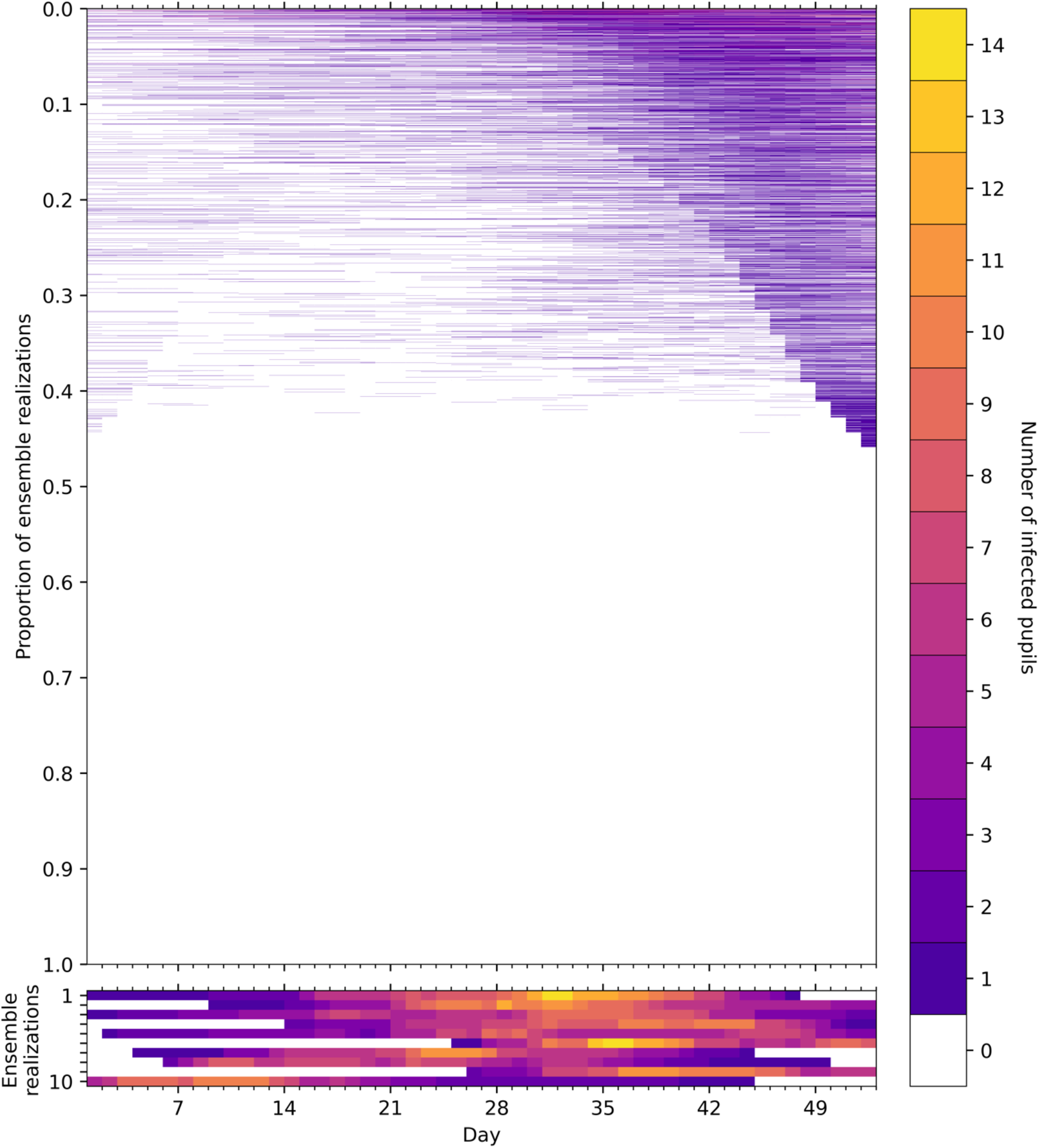
Time series of the number of infected pupils for each realization in an ensemble of 100,000 stochastic simulations of an isolated classroom in Term 1 of 2020. Each row of the heatmap corresponds to an ensemble member, with colours denoting the number of pupils in the classroom with Covid-19 infection on each day. The ensemble realizations have been sorted by the sum of the number of infected pupils over all days. (Upper panel) All ensemble realizations are shown. (Lower panel) The ten ensemble realizations with the most infections are shown, illustrating the dynamics of large outbreaks that occur with a frequency of 1/10,000. Colours denote the number of infected pupils in the classroom for each day of the simulation.

**Figure 7.**
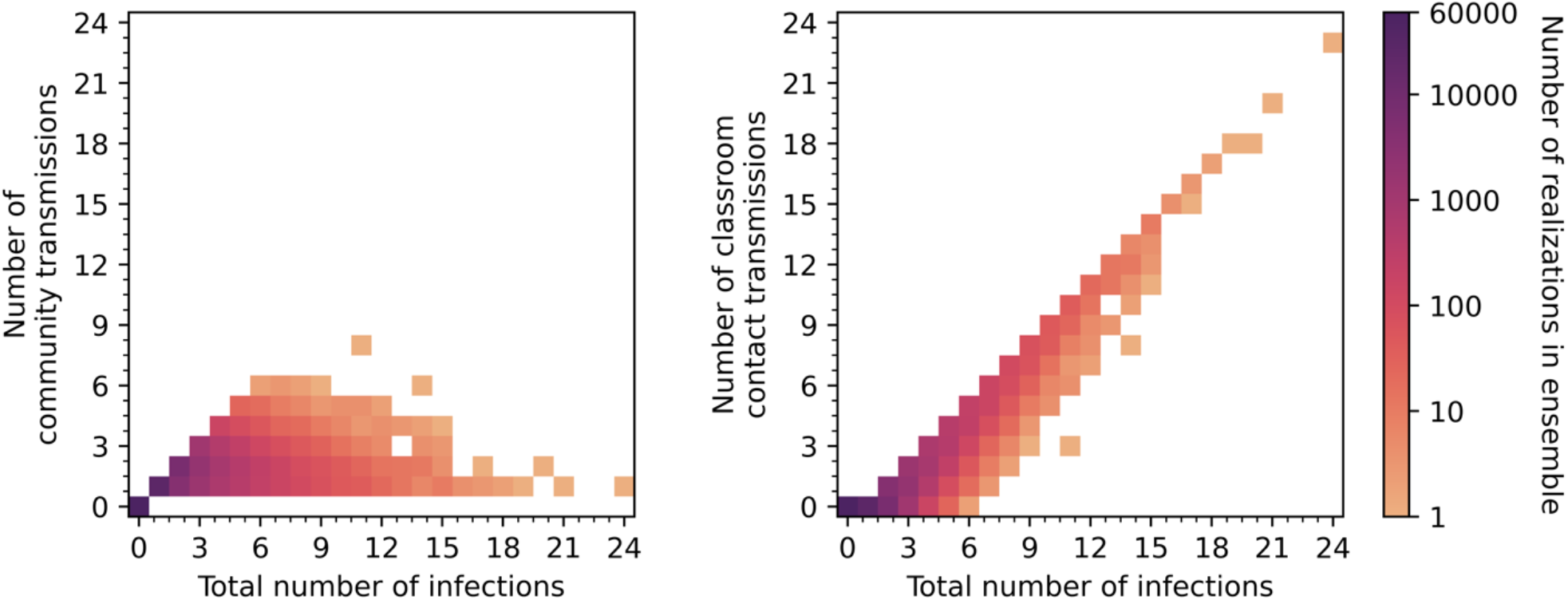
The number of infections caused by community transmissions and classroom contact transmissions in the ensemble simulations as functions of the total number of infections. The intensity of colour of the points indicates the frequency of occurrence within the ensemble on a logarithmic scale, and the ensemble has 100,000 members.. Note, stochastic variations are apparent for low frequencies.

When the total number of infections is up to six, we find that most infection occurs due to community transmission (Figure 7a), but beyond six infections this correlation is broken and in all these simulations (with infections >5) the number of classroom infections significantly exceeds the number of infections expected in the community. These cases could pragmatically be defined as ‘clusters’ or ‘outbreaks’ due to in-class transmission being dominant. Current Department for Education (DfE) contingency operational guidance for schools [30] defines two thresholds for “extra action” under specific setting conditions and in relation to ‘close mixing’ groups within schools: “5 children, pupils, students or staff, who are likely to have mixed closely, test positive for COVID-19 within a 10-day period; or 10% of children, pupils, students or staff who are likely to have mixed closely test positive for COVID-19 within a 10-day period”. These criteria relate to two completed rounds of tests in an ‘asymptomatic test sites’ regime, initiated only following full return to school; it is not clear if there are criteria for similar actions due to in-school outbreaks under other circumstances.

In our ensemble there are relatively few realizations (approximately 1%) with total infections exceeding six, suggesting such outbreaks are relatively uncommon. Larger outbreaks of ten infected pupils, do not occur in our ensemble unless classroom transmission occurs. We find outbreaks with ten or more infected pupils occur in ∼0.3% of the simulations, while very large outbreaks of 15 or more pupils (i.e., at least half of our simulated class are infected) occur in only 25 (0.025%) of the realizations.

Open data on which to compare our results is scarce, but some summary statistics on school attendance have been published through the pandemic by the UK Department for Education (DfE) [31]. These statistics report that on 22 October 2020, about 20% of state-funded primary schools reported one or more pupils who were self-isolating. While it is difficult to compare this summary statistic with our stochastic simulations, as we do not know how many classes had pupils in self-isolation, the value compares favourably with our ensemble which has 18,028 (18%) of the realizations having one or more pupils quarantined on this day of the simulation. Based on the simulations we also expect about 2665 outbreaks.

### Assessing effectiveness of mitigation approaches

To examine the effectiveness of mitigation approaches, we perform stochastic ensemble simulations with different infection control measures imposed. We model a collection of 10 primary schools, each consisting of four classrooms with 30 pupils and one teacher. The Term 1 variation in community prevalence and incidence rate is adopted to model the changes over time in the external community epidemic and allows us to provide a comparison with the simulations above where there are no hard mitigation measures included.

In Figure 8 we compare the effect of the contact rates in classrooms and the mitigation approaches on the total number of pupils infected over the Term1 period within the 40 classrooms consisting of 1200 pupils in total. The box plots show the median, 25^th^ and 75^th^ percentiles, with the interquartile range (IQR) being their difference, the lower (25^th^ percentile − 1.5 × IQR) and upper (75^th^ percentile + 1.5 × IQR) Tukey fences, and outliers.

**Figure 8.**
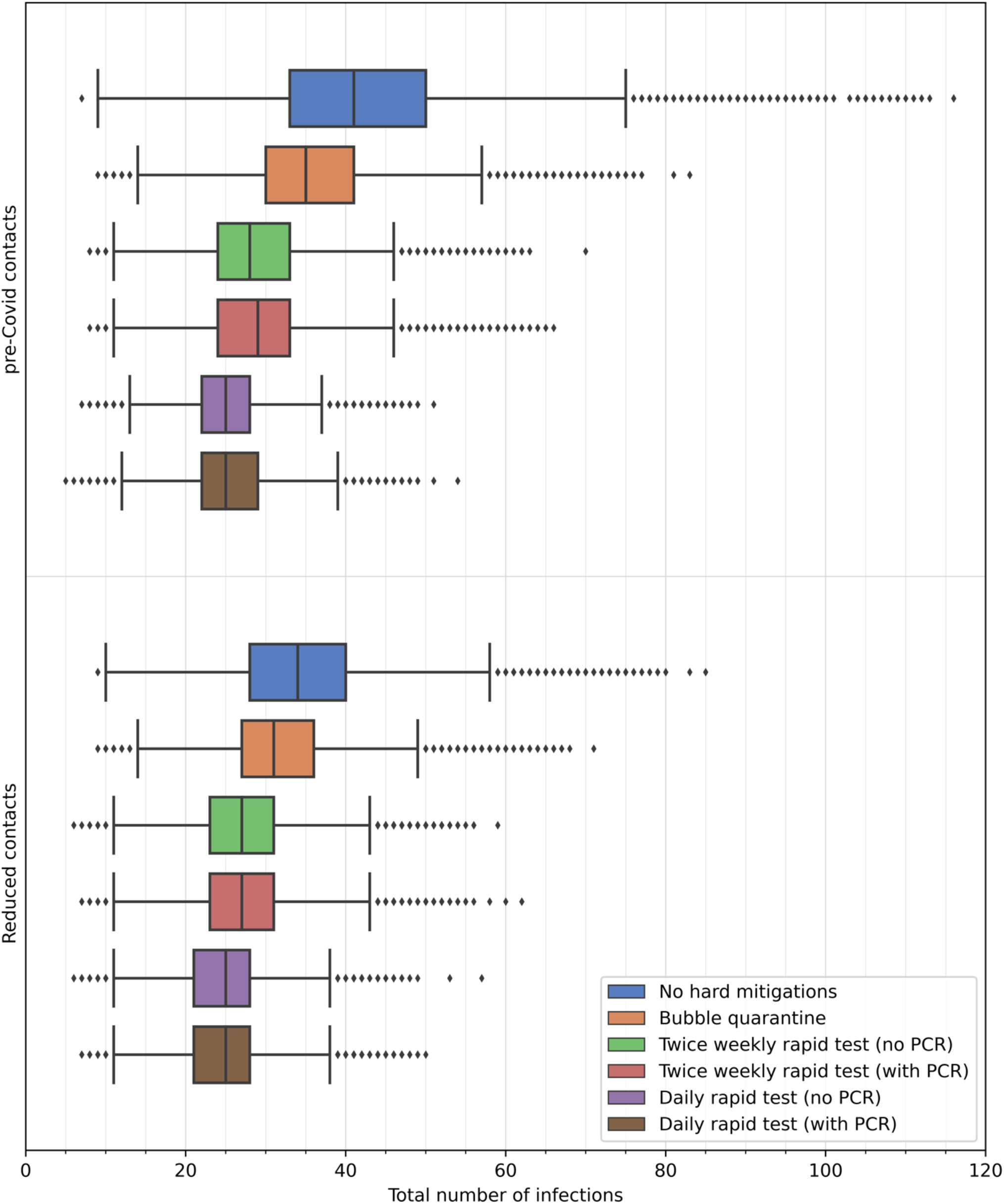
Comparison of the number of infected pupils in 40 primary classrooms with different mitigation measures applied. The total number includes those initially infected and those infected from community transmission. Box and whisker plots illustrate median, interquartile range, Tukey fences and outliers. Contact rate distributions for pre-Covid times and in small classes are derived from SEJ and used to construct distributions of expected contact rates for schools during Term 1 in September—October 2020. Mitigation measures simulated are: (i) ‘No hard mitigations’ where only individuals who are unwell are quarantined (blue); (ii) ‘Bubble quarantine’ where an unwell individual results in quarantine of the whole class (orange); (iii) ‘Twice weekly rapid test (no PCR)’ where each individual is tested with a rapid lateral flow test twice each week, with positive test results resulting in quarantine of the individual (green); (iv) ‘Twice weekly rapid test (with PCR)’ where each individual is tested with a rapid lateral flow test twice each week, with positive test results resulting in quarantine of the individual and a confirmatory PCR test is applied after a two-day interval (red); (v) ‘Daily rapid test (no PCR)’ where each individual is tested with a rapid lateral flow test daily, with positive test results resulting in quarantine of the individual (purple); (vi) ‘Daily rapid test (with PCR)’ where each individual is tested with a rapid lateral flow test daily, with positive test results resulting in quarantine of the individual and a confirmatory PCR test is applied after a two-day interval (brown).

We consider first the simulations where no ‘hard’ mitigation measures are applied (blue boxes in Figure 8). Here, the contacts rate distributions have a strong influence on the number of infections. With the contact rates of pre-Covid classrooms, the median number of pupils infected is 41 (75^th^ percentile: 50). With reduced contacts, the median number of infections is reduced by 17% to 34, with decreases also in the spread of values, particularly on the upper tail (75^th^ percentile: 40).

Each of the mitigation measures reduces the number of infected pupils, but they are generally less effective than the changes in the contact rates. Where contact rates are highest (i.e., using the pre-Covid contact rate distributions) the mitigation measures each substantially reduce the number of infections, as seen in the decrease in the quartiles, Tukey fences and outliers. The extent of the decrease in these statistics diminishes as contact rates decrease.

We compare each of the mitigation measures using the reduced contacts rates adopted for primary schools in Term 1 of 2020. Bubble quarantine lowers the upper quartile and upper fence values but does not substantially change the median number of infected pupils across the ensemble (Figure 8), lowering the median slightly from 34 to 31. This result is expected as bubble quarantine is not very effective in guarding against the seeding of infection carried into the classrooms from the community, and the spreading of this infection in the incubation period before an infected individual has symptoms that trigger the quarantine period. However, the reduction of the upper tail of the distribution (Figure 9) shows that bubble quarantine can produce a decrease in the occurrence of outbreaks. For example, small outbreaks of six of more infected pupils occur in 0.6<u>0</u>% of isolated classroom ensemble realizations (reduced from 1.7% with no hard mitigations), outbreaks of ten or more occur in 0.03% of realizations (c.f. 0.3% with no hard mitigations), and large outbreaks of 15 or more infections occur in only 0.002% of the realizations (c.f. 0.025% with no hard mitigations) (Table 2).

**Table 2.** Proportion of realizations in ensemble of 100,000 isolated classroom simulations that produce outbreak with more than six infected pupils, large outbreaks with ten or more infected pupils, and very large outbreaks with 15 or more infected pupils. Six mitigation measures are simulated, as detailed in the main text. Zero value indicate no realizations occur in the ensemble. Simulations were conducted using the reduced contact rates distribution.

| Mitigation measure | Proportion of outbreaks (>6 infected) | Proportion of large outbreaks ($\geq 10$ infected) | Proportion of very large outbreaks ($\geq 15$ infected) |
| --- | --- | --- | --- |
| No hard mitigations | 1.06% | 0.28% | 0.025% |
| Bubble quarantine | 0.28% | 0.028% | 0.002% |
| Twice weekly rapid test (no PCR) | 0.08% | 0.007% | 0 |
| Twice weekly rapid test (with PCR) | 0.093% | 0.01% | 0 |
| Daily rapid test (no PCR) | 0.003% | 0 | 0 |
| Daily rapid test (with PCR) | 0.007% | 0 | 0 |

**Figure 9.**
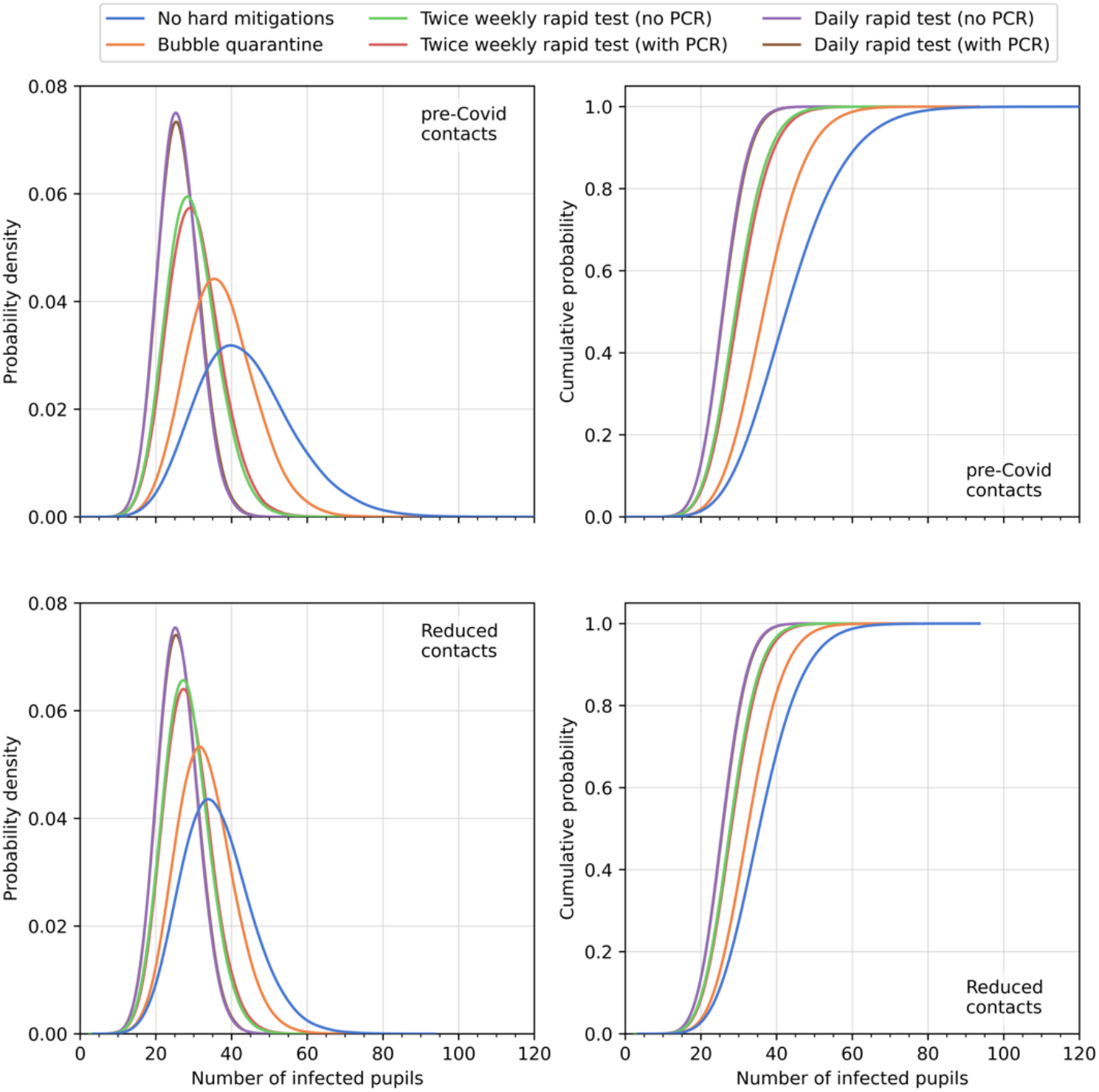
Comparison of the number of infected pupils in 40 primary classrooms with different mitigation measures applied. Estimates of the cumulative distribution functions (left) and probability density functions (right) using Kernel Density Estimation for each of the mitigation strategies are shown.

Considering next the use of rapid testing, we simulate approaches where each member of the classrooms is tested at regular intervals. Specifically, we model a twice weekly testing regime, such as that used in summer 2021, and a daily test regime. Further, we simulate each with and without confirmatory PCR testing, with a two-day time lag for conducting the PCR test.

With twice weekly testing without PCR testing, there is a decrease in the median number of infections to 27, a narrowing of the interquartile range with a pronounced reduction in the upper quartile to 31 infections, and a reduction in the upper fence value to 43 infections (Figure 8). This suggests that a test-based surveillance approach is effective in reducing infections within school through the disruption of contact networks. Large outbreaks, in particular, are less common, occurring in approximately 0.007% of ensemble realizations (Table 2), and very large outbreak did not occur in the simulations. Confirmatory PCR testing has minor effects on the distribution of the number of infections, increasing only the maximum value in this sample to 62 (noting that the outliers are rare events and fluctuate when resampling). This result can be rationalized as there is a small non-zero probability of a ‘false negative’ PCR test overturning a ‘true positive’ rapid test and thus returning an infectious individual into the class. The frequency of outbreaks slightly increases in the ensemble.

Implementing daily rapid lateral flow testing frequency further shifts the distribution of the number of infections to lower values (Figure 8 and Figure 9), although the change is modest (median: 25; 75^th^-percentile: 28 for simulations both with and without PCR testing), representing a change of only 7% in the median and 10% in the upper quartile values compared to the twice-weekly rapid testing ensembles. Outbreaks are further reduced (albeit from small values for twice-weekly testing), and no large outbreaks of ten or more infected pupils occur in the ensemble (Table 2). In this case the effect of confirmatory PCR testing is barely discernible in the numbers of infected pupils, with only far outliers differing (the interpretation of which should be made cautiously without larger ensembles). This can also be explained: daily testing ensures that ‘false negative’ test results are rapidly re-tested, and sequences of repeated false negatives are unlikely even for the low sensitivity rapid test. There is an increase in the number of outbreaks within the ensemble, although the proportion remains small.

Where infection occurs in the ensemble realizations, we track the number of secondary infections within the classroom from each infected individual over the duration of the simulation. The mitigation measures shrink the number of secondary infections by reducing contact transmission. This is illustrated in Figure 10 which shows box and whisker plots for the proportion of infections that occur due to classroom transmissions. The reduction of contact rates, without additional mitigation measures, has a substantial effect on the proportion of classroom transmissions, reducing the median from 0.41 with pre-Covid contacts to 0.28 with reduced contacts (a ∼30% reduction), and similar relative reduction in the quartile values. The mitigation measures further reduce the proportion of classroom transmissions, with the testing-based approaches having greater effect than the bubble quarantine. Indeed, with reduced contacts rates and daily rapid testing, the median proportion of classroom transmissions is zero, and the 99^th^ percentile value is 0.12 when there is no confirmatory PCR testing (which increases to 0.15 when PCR testing is applied).

**Figure 10.**
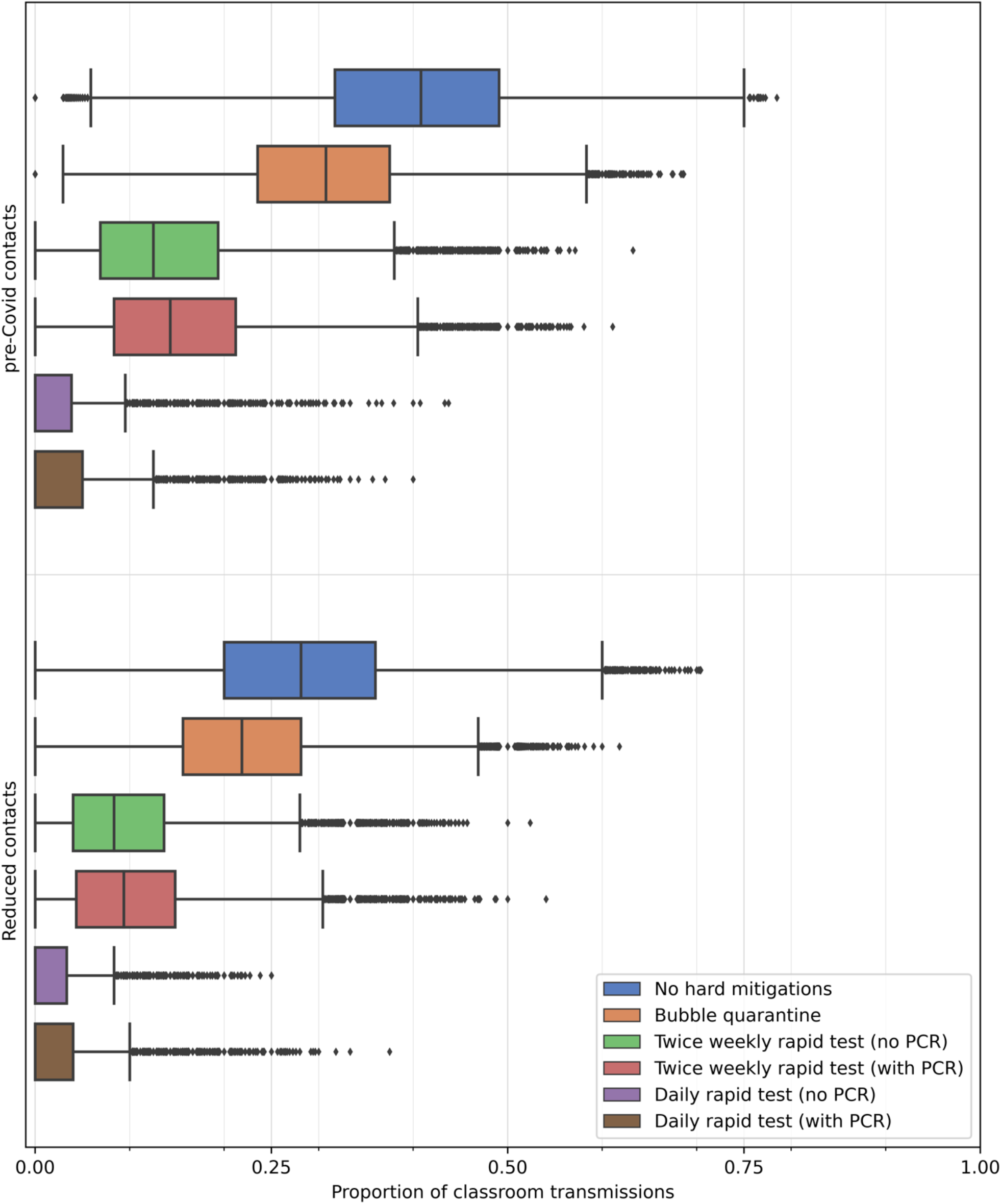
Box and whisker plots comparing the proportion of infections resulting from classroom transmission in 40 primary classrooms with different mitigation measures applied in schools during Term 1 in September—October 2020. Box and whisker plots illustrate median, interquartile range, Tukey fences and outliers. Contact rate distributions for and mitigation measures are as for Figure 8.

The ensemble results can be used to estimate the secondary infection rate in the classrom as the expected number of in-classroom infection transmissions. Figure 11 shows two summary statistics (the mean and 99^th^-percentile values) to characterize the distribution of the number of secondary infections that occur in the ensemble simulations. To highlight the trends when curtailing contact rates, for Figure 11 have conducted simulations here with the contact rate distributions derived from SEJ for small classes and applied these rates to full classroom attendance; it is unlikely that such low contact rates could be maintained in a fully occupied classroom, but the simulations are illustrative hypothetical scenarios.

**Figure 11.**
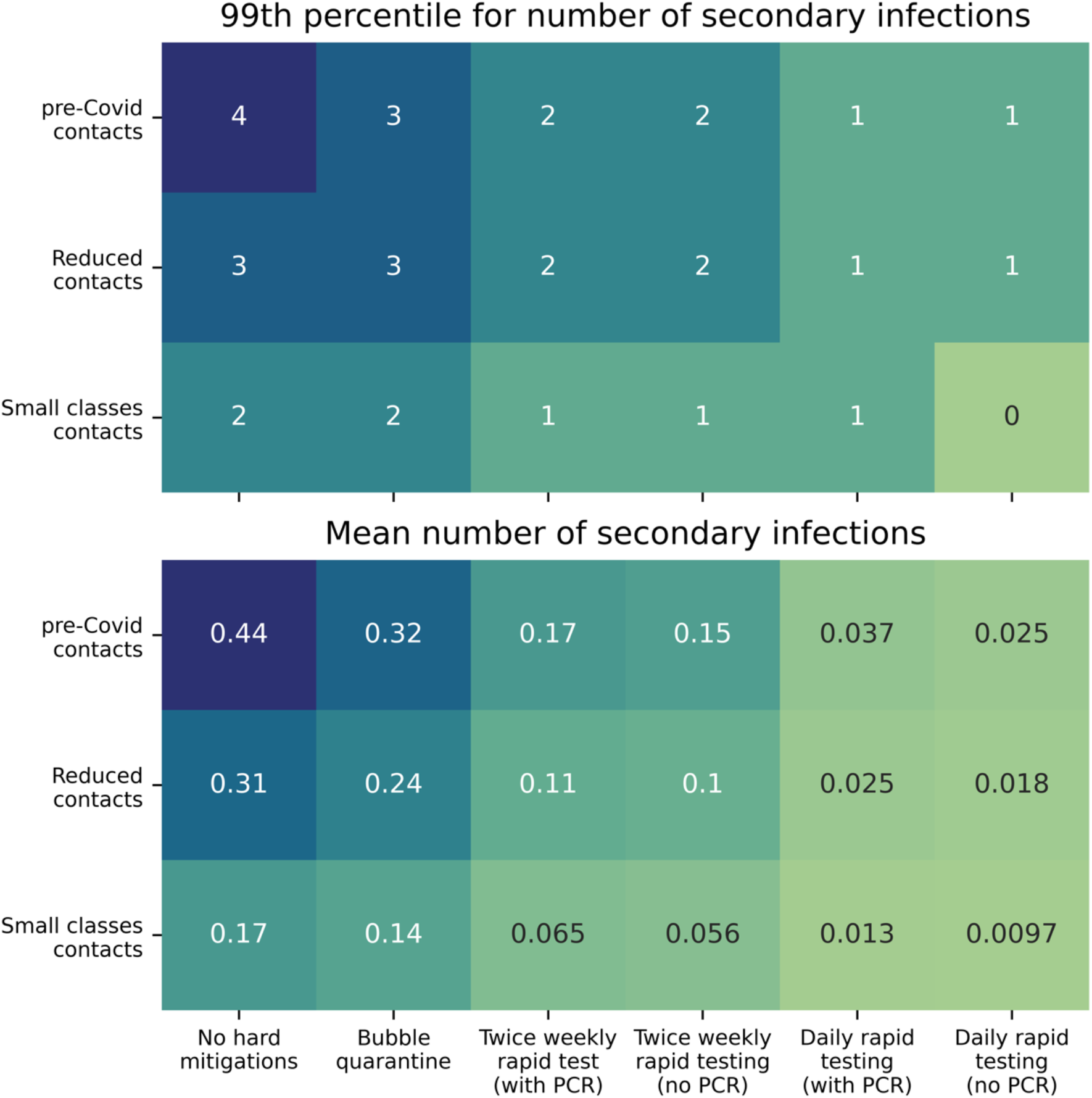
The number of secondary infections in classroom simulations with different contact rates distributions and mitigation measures applied. Two summary statistics to characterize the distributions are shown, with the 99^th^ percentile value (upper panel) and mean value (lower panel) of the number of secondary infections in all 100,000 ensemble simulations calculated. ‘Small classes contacts’ refers to simulations that adopt contact rate distributions derived from SEJ for small classes that are applied to full classroom attendance. Note, most of infections in the simulations are not transmitted in classroom contacts, so mean values are below 1.0 in all cases.

The mean value of the number of secondary infections is less than unity in all cases, indicating that the infection is not transmitted in the classroom contacts of most primary cases. Each mitigation measure reduces the mean value of the number of secondary infections, with more pronounced effects when the contact rate is high.

Outbreaks in the classroom can be driven by relatively rare ‘*super-spreaders*’. Our model includes the influence of having some very gregarious individuals in a mixing group, as reported in the contact in [2]. Here we illustrate the impact of mitigation measures on super-spreaders by calculating the 99^th^ percentile value for the number of secondary infections (i.e., 1% of infected individuals in the simulations have a secondary infection number larger than the 99^th^ percentile value). The mitigation measures are again seen to reduce the 99^th^ percentile values, although the ‘bubble quarantine’ approach has only modest (if any) affect. The testing-based surveillance approaches are more effective in eliminating super-spreading, reducing the 99^th^ percentile value of the number of secondary infections to unity (or smaller) if daily testing is implemented.

The mitigation measures have consequences on pupil absences. The ‘bubble quarantine’ is particularly severe, with whole classes being quarantined for a period of 10 days if there is a single symptomatic case. However, the testing-based surveillance approaches leads to self-isolation of individual cases, so the impact on absences is greatly reduced. In Figure 12 we illustrate the effects of the mitigation measures on the number of pupils present in the classroom on one day, 23 October 2020 which we took as the last day of Term 1 in 2020. As the majority of the realizations in the ensemble have no infections, there is typically full attendance in each of the scenarios simulated. However, when ‘bubble quarantine’ is employed, there is full-class quarantine on this day in greater than 15% of the ensemble members. If this is result is applied to all classrooms in England, this would suggest approximately 25,000 classrooms in quarantine on this day and, with an average class size of 27 in England [27], approximately 675,000 pupils absent from school. For the testing-based surveillance, there are fewer absences, with only ∼0.1% of the ensemble simulations having more than five pupils in quarantine on the last day of the school term. The contact rate distribution is again seen to play a prominent role, with reduced contacts leading to reduced absences as there are fewer infections to trigger quarantine.

**Figure 12.**
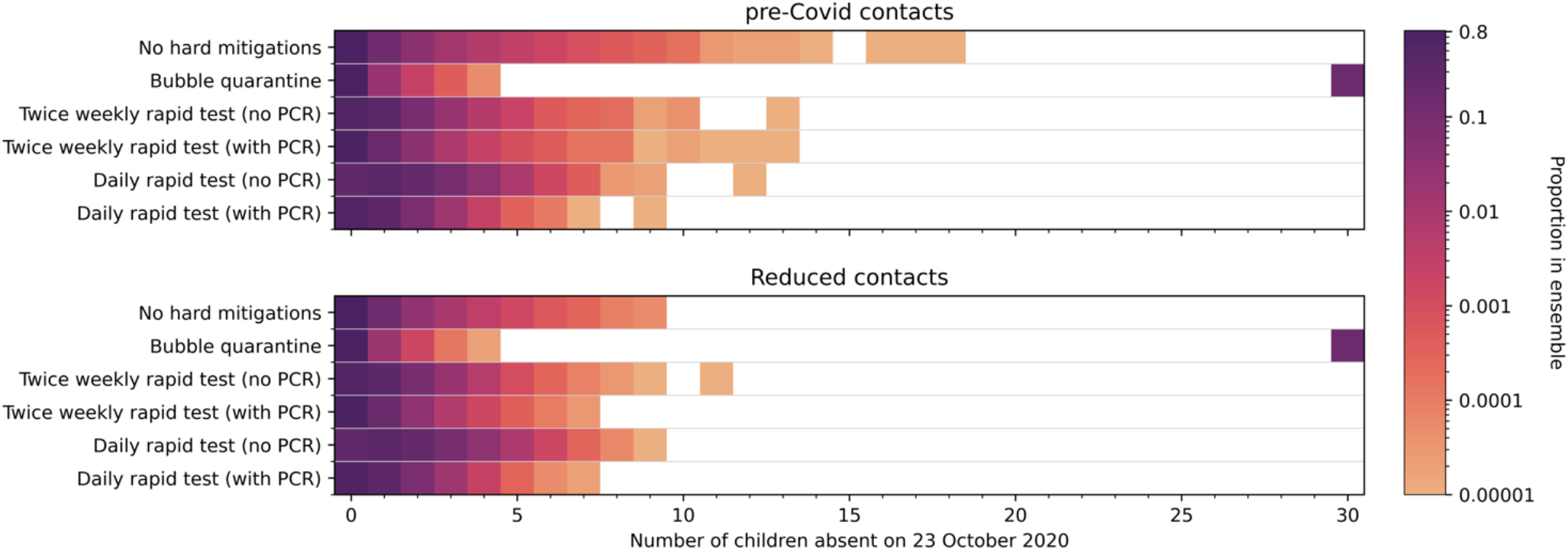
Number of pupils absent from school on 23 October 2020 (the last day of Term 1 in 2020) in an ensemble of 100,000 simulations of a class of 30 pupils. Two contact rates are applied: pre-Covid contact rates (top panel) and reduced contact rates (bottom panel). Six mitigation measures are modelled. Colours indicate the proportion of ensemble realizations on a logarithmic scale (white indicating no occurrence in the ensemble). Apparent gaps in some of the rows occur due to the probability events which may not occur in the finite ensemble.

To assess the impact of the testing strategies, we focus here on the ‘Reduced contacts’ distribution (lower panel in Figure 12). Twice weekly rapid testing has similar proportions absent from the classroom as the simulations with no hard mitigations applied. Without confirmatory PCR testing, twice weekly rapid testing slightly increases the number of ensemble members for which there is a single pupil absent, while the application of a confirmatory PCR test returns this proportion to close to that found when no hard mitigations measures are applied. In contrast, daily testing without PCR confirmation has a substantially elevated proportion of simulations with classroom absences, with more than ∼1% of simulations having five pupils absent. Confirmatory PCR testing reduces the absentee proportions to below that when no mitigation measures are applied. This confirms that many of those quarantined by daily rapid testing are done so unnecessarily and are promptly returned to the classroom when a PCR test overturns a false-positive rapid test. However, the disruption to education by missing some days at school is significant and the benefit from a public health perspective is negligible.

We estimate the numbers absent on a national scale by performing a bootstrap resampling of our ensemble to produce a sample of 156,843 classes, matching the number of state primary schools in 2020—2021 [27]. As our nominal class size is 30 pupils, this gives a total number of pupils of 4,705,290 which is comparable to the national state-primary pupil count of 4,177,058 [27] (a different of -13% in the simulated classes). The bootstrap ensemble is recomputed 10,000 times to construct a median value for the number of pupils absent on 23 October 2020 and confidence intervals on our estimates, which are reported in Table 3.

**Table 3.** Estimated national number of pupils absent from school on 23 October 2020 from ensemble simulations with bootstrap resampling. Isolated classes are simulated with six mitigation strategies and two contact rate distributions. Ensembles of 100,000 classes are simulated and collections of 156,843 classes are formed by bootstrap resampling from the ensembles. Each bootstrap is constructed 10,000 times to obtain distributions on the number of absences, and we report the median, 5^th^ and 95^th^ percentile values. The percentage of the school pupil population is estimated assuming each class has 30 pupils, giving a school population of 4,705,290.

|  | Mitigation measure | Number of pupils absent |  | Percentage of pupils absent |  |
| --- | --- | --- | --- | --- | --- |
|  |  | median | 5<sup>th</sup>; 95<sup>th</sup> percentiles | median | 5<sup>th</sup>; 95<sup>th</sup> percentiles |
| Pre-Covid contacts | No hard mitigation | 48,384 | 47,861; 48,908 | 1.03 | 1.02; 1.04 |
|  | Bubble quarantine | 762,605 | 755,422; 769,729 | 16.21 | 16.05; 16.35 |
|  | Twice weekly rapid test (no PCR) | 95,529 | 94,977; 96,069 | 2.03 | 2.02; 2.04 |
|  | Twice weekly rapid test (with PCR) | 45,995 | 45,592; 46,416 | 0.98 | 0.97; 0.99 |
|  | Daily rapid test (no PCR) | 202,887 | 202,152; 203,617 | 4.31 | 4.30; 4.33 |
|  | Daily rapid test (with PCR) | 81,329 | 80,855; 81,811 | 1.73 | 1.72; 1.74 |
| Reduced contacts | No hard mitigation | 40,734 | 40,312; 41,174 | 0.87 | 0.86; 0.88 |
|  | Bubble quarantine | 748,469 | 741,461; 755,625 | 15.9 | 15.8; 16.1 |
|  | Twice weekly rapid test (no PCR) | 93,517 | 92,980; 94,053 | 1.99 | 1.98; 2.00 |
|  | Twice weekly rapid test (with PCR) | 42,991 | 42,615; 43,374 | 0.91 | 0.91; 0.92 |
|  | Daily rapid test (no PCR) | 203,566 | 202,836; 204,305 | 4.35 | 4.31; 4.34 |
|  | Daily rapid test (with PCR) | 80,725 | 80,248; 81,199 | 1.72 | 1.71; 1.73 |

Table 3 shows that the different mitigation strategies have a strong effect on school absence, with the model adopting bubble quarantine resulting in more than 15% of pupils absent from school on 23 October 2020. The use of rapid testing reduces the numbers of absent pupils substantially, decreasing the median value to 2% of the national primary school pupil population for twice weekly testing with no PCR confirmation. This is further decreased to around 1% of the pupil population if confirmatory PCR testing is used to return pupils with false-positive rapid test results to the classroom. However, daily rapid testing produces many more false-positive results, so that the number of absent pupils increased markedly to over 200,000 (approximately 4% of the pupil population) when there is no confirmatory follow-up PCR test. With a PCR test to identify false-positive cases of daily rapid tests, the number of absent pupils decreases to approximately 80,000 (1.7% of the pupil population) which is comparable to the number absent with twice-weekly rapid testing with no PCR follow-up.

## 5. Forecasting SARS-CoV-2 in schools in 2021

In August 2021, the safe return of pupils to schools was a national concern, with increasing prevalence over the summer holidays and emerging predominance of the more-transmissibly Delta variant. To estimate SARS-CoV-2 infection transmission and assess the effectiveness of mitigation measures, we performed simulations in a ‘forecasting’ capacity in August 2021, applying our model to the first seven weeks of the new school year (i.e., assumed here to be Thursday 2 September to Friday 22 October 2021 of Term 1).

### Amendments to the model

To apply our model to forecasting SARS-CoV-2 in Term 1 of 2021, we alter our model parameters from those used for Term 1 2020. In particular, we include the impact of population vaccination by September 2021. Here we assume that there is 95% uptake amongst classroom staff, and that 1% of children have been vaccinated.

Another crucial difference is the high prevalence of the Delta (B.1.617.2) variant in the UK population. Genomic surveillance [32] and modelling studies [19-21] indicate that the Delta variant is expected to be predominant in the UK in September 2021, accounting for more than 95% of Covid-19 infection cases [19-21].

The Delta variant has two major impacts on our updated model. Firstly, it has substantial transmission advantage over the original wildtype SARS-CoV-2 [33, 34]. This is estimated as 61% transmission advantage over the Alpha (B.1.1.7) variant in [33], from which we estimate as a transmission advantage of 140% over the original wildtype. We therefore increase our values of the probability of contact transmission from original wildtype SARS-CoV-2 values using this scaling factor. The probabilities of contact transmission we adopt for the Delta variant are given in Table 4 (again assuming, as before, that adults are twice as likely as children to transmit infection [6-8]).

**Table 4.** Probability of transmission of Delta variant SARS-CoV-2.

| Delta-variant transmission probability | Adults | Children |
| --- | --- | --- |
| Symptomatic | 0.084 | 0.042 |
| Asymptomatic | 0.059 | 0.029 |

An additional characteristic of the Delta variant, which drives increased infection, is a shortening of the incubation period for infected individuals [35]. Delta-variant infection is detectable (by PCR testing) in cases approximately two-days earlier than for original wildtype SARS-CoV-2 infection [35]. We adopt this shorter interval in our model by reducing the median duration of the incubation period by two days. Thus, for each individual in our population, we assign an incubation time drawn from log-Normal distribution [5] with *t*_*i*_ ∼log*N*(1.10, 0.5). In principle, the variance of the log-Normal distribution could also be amended, but in the absence of detailed information, we retain the previous value. The reduced incubation period affects the time-dependence of the relative transmission probability, with a more rapid increase to peak-infectiousness for the Delta-variant model parameters. Therefore, the probability of infection transmission from pre-symptomatic individuals is enhanced when considering the Delta-variant.

### Forecast scenarios

At the time our forecast simulations were performed there was significant uncertainty in the trajectory of the epidemic into the new school year. Projections from national-scale epidemiological models fitted to observations [19-21] provide a basis for forecast scenarios. Typically, the projections in [19-21] suggested a third wave of the epidemic was likely in late-summer as the effects of easing of restrictions on 19 July on infection transmission were realized. The model projections also indicated that infection amongst younger age-groups would become dominant [19]. The magnitude of a third wave varies across models and with model assumptions, particularly regarding social mixing over the summer, and peak incidence rates (per 10,000 population per day) range from ∼12 [19] to ∼300 [20] across the models, spanning values substantially higher than those for September 2020 (Figure 5; [18]).

To model the community incidence rate of new infections over Term 1 of 2021, we digitize projection graphics produced by a national scale epidemiological model in [19] (Figure 13). This provides two time series of incidence rate which differ in the magnitude and timing of the peak depending on how rapidly societal contacts returned to pre-Covid levels which we refer to as the ‘gradual-relaxation’ and ‘rapid-relaxation’ incidence rates (Figure 13). For the gradual-relaxation scenario, a peak incidence rate of approximately 67,000 new infections per day was projected to occur in late July, with a gradual decrease through August and September, to approximately 20,000 per day on 1 October 2021, with incidence slightly increasing subsequently [19]. In this scenario, our simulations of the school term begin in a period of relatively steady incidence. In contrast, if societal contact rates rapidly return to pre-pandemic levels in summer 2021, then the projected peak incidence rate was approximately 145,000 new infections per day and occurs on 12 August 2021, with incidence rates elevated with respect to the gradual-relaxation until 1 October 2021, from when the incidence rate gradually decreases to low levels through October and November [19]. The incidence of new infection was not expected to be evenly distributed through the population, but rather skewed to lower age groups who were unvaccinated. Here, we adopt the values reported in [19] that attribute 27% and 25% of new infections to age groups 0—9 and 10—19, respectively, for the gradual-relaxation scenario, and combine these to produce a child-incidence rate. These proportions change but slightly for the rapid-relaxation scenario to 26% and 24% for age groups 0—9 and 10—19, respectively.

**Figure 13.**
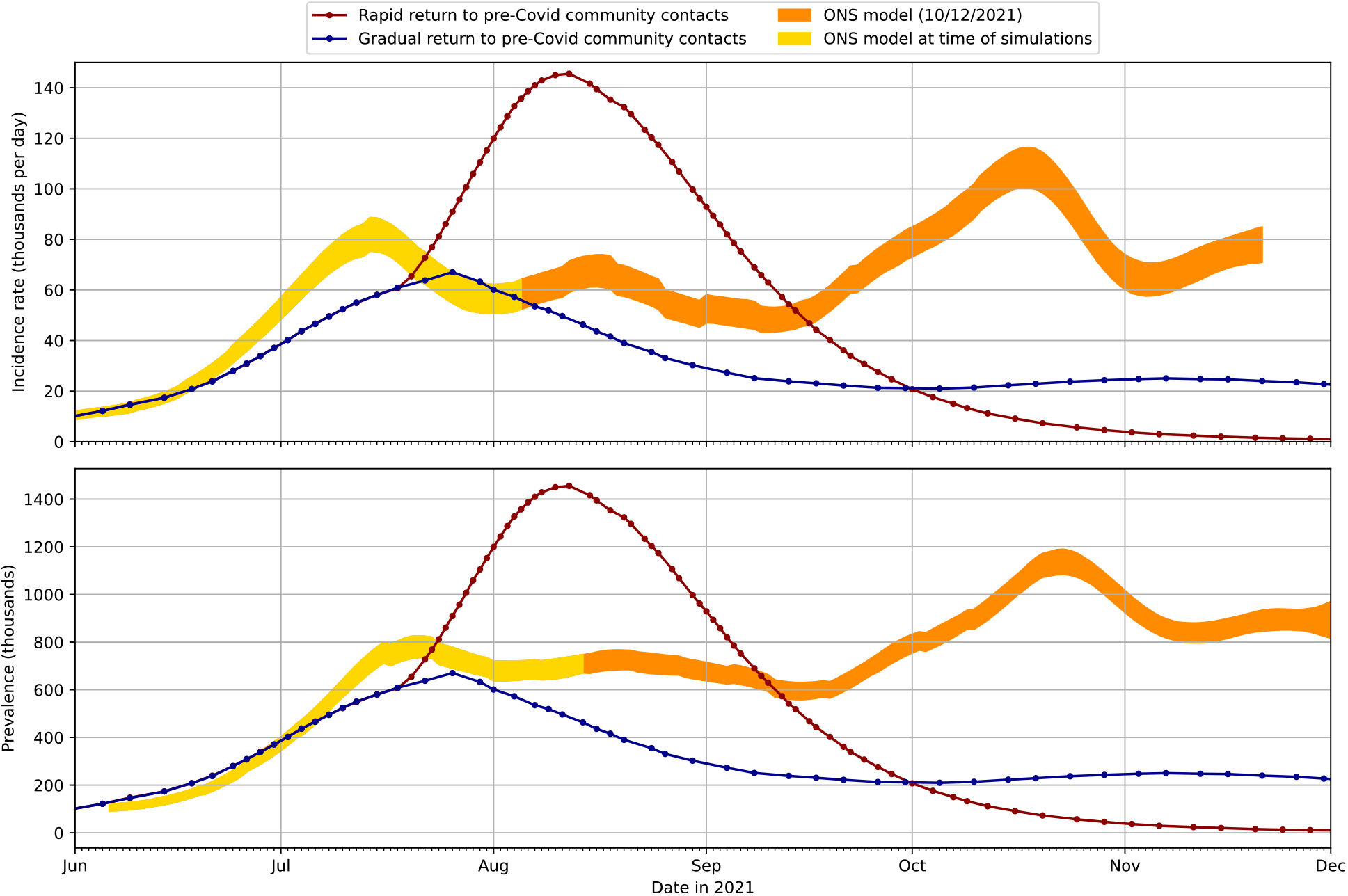
Projections of the incidence rate (upper panel) and prevalence (lower panel) in the UK from June to December 2021. Incidence rates are obtained from a national scale epidemiological model [19] with scenarios modelling a gradual or rapid return to pre-pandemic societal contact rates. The prevalence is modelled assuming R_F_ = 5 and infection duration of 40 days (see supplementary material). Data from the Office for National Statistics Coronavirus Infection Survey [29] for the modelled incidence and prevalence with credible intervals are illustrated for the data available at the time of writing (early December 2021, orange) and the data available at the time simulations were conducted (late-August 2021, yellow).

At the time the forecast simulations were performed (mid-August 2021), the projections of community incidence rate were tracking well the estimates derived from the national Coronavirus Infection Survey compiled by the Office for National Statistics [29] (noting that the national scale projections were made in early July 2021), with the gradual-relaxation scenario in particular predicting quite well the timing and magnitude of the early summer ‘peak’ in incidence. Subsequently, however, observed incidence rates did not fall as substantially as the projections anticipated, and there are significant and increasing deviations in the projectioned incidence rate from observations from mid-September and into October. Therefore, as our classroom transmission model adopts the projected incidence rate for community-transmission seeding of infection into schools, we expect our forecasts should differ from observed infection rates in schools.

Our model also requires the community prevalence. This is not reported in the projections, so we estimate prevalence using a simple rescaling of incidence rates with comparison to the available data (Figure 13), as described in the supplementary material Appendix A3. The prevalence has less importance in our model than the incidence rate (as it is used only as a seeding of infection on model initiation and in cases where substitute teachers are added to the simulated classroom) and we judge that the estimate from incidence rate is sufficient in our forecasts. However, we note that, as with the incidence rate, from mid-September there are substantial deviations of the projection-derived prevalence from those observed subsequently observed.

Guidance from the Department for Education issued in summer 2021 [36] removed many of the SARS-CoV-2 mitigation measures in school settings, but recommends ‘soft mitigation’ measures continue to be applied, such as enhanced cleaning and classroom ventilation. Therefore, in our forecast simulations, we do not model any of the possible hard mitigations that might be activated under certain conditions. Additionally, schools were able to reconvene in large groups, such as during lunch and break times and for school assemblies. This may increase transmission rates within school settings and particularly between classes. However, school managers were responsible for deciding whether to continue to ‘bubble’ classes to minimize interactions. As teachers and school leaders could also decide to continue to implement measures that reduce contacts within the classroom, we adopt the two contact rates distributions from [2] that model contact rates for pre-Covid classrooms and reduced contact rates during the epidemic.

### Classroom transmission modelling results

We perform stochastic simulations for an isolated school classroom over the 50 days of Term 1 in 2021, adopting the ‘reduced contacts’ distribution of contact rates (i.e., we assume that in this time period, schools will continue to enact measures to reduce the number of contacts in classrooms).

Figure 14 illustrates the result when for the ‘gradual-relaxation’ scenario, showing time series of the number of infected pupils in each realization from an ensemble of 100,000 stochastic simulations in which no hard mitigation measures are applied. Therefore, Figure 14 is directly comparable to Figure 6, and shows there is potential for substantially increased infection within school classrooms in Term 1 of 2021 when compared with Term 1 of 2020. We find both (i) a potential substantial increase in the proportion of ensemble realizations that have infected pupils (62% for 2021, compared to 46% for 2020) and (ii) the maximum number of simultaneously infected pupils in a realization could be increased (22 in 2021, compared to 14 in 2020). There are also more realizations in the ensemble that have several simultaneously infected pupils, although most frequently there is only a single infected pupils in the class, as we found in the Term 1 2020 simulations.

**Figure 14.**
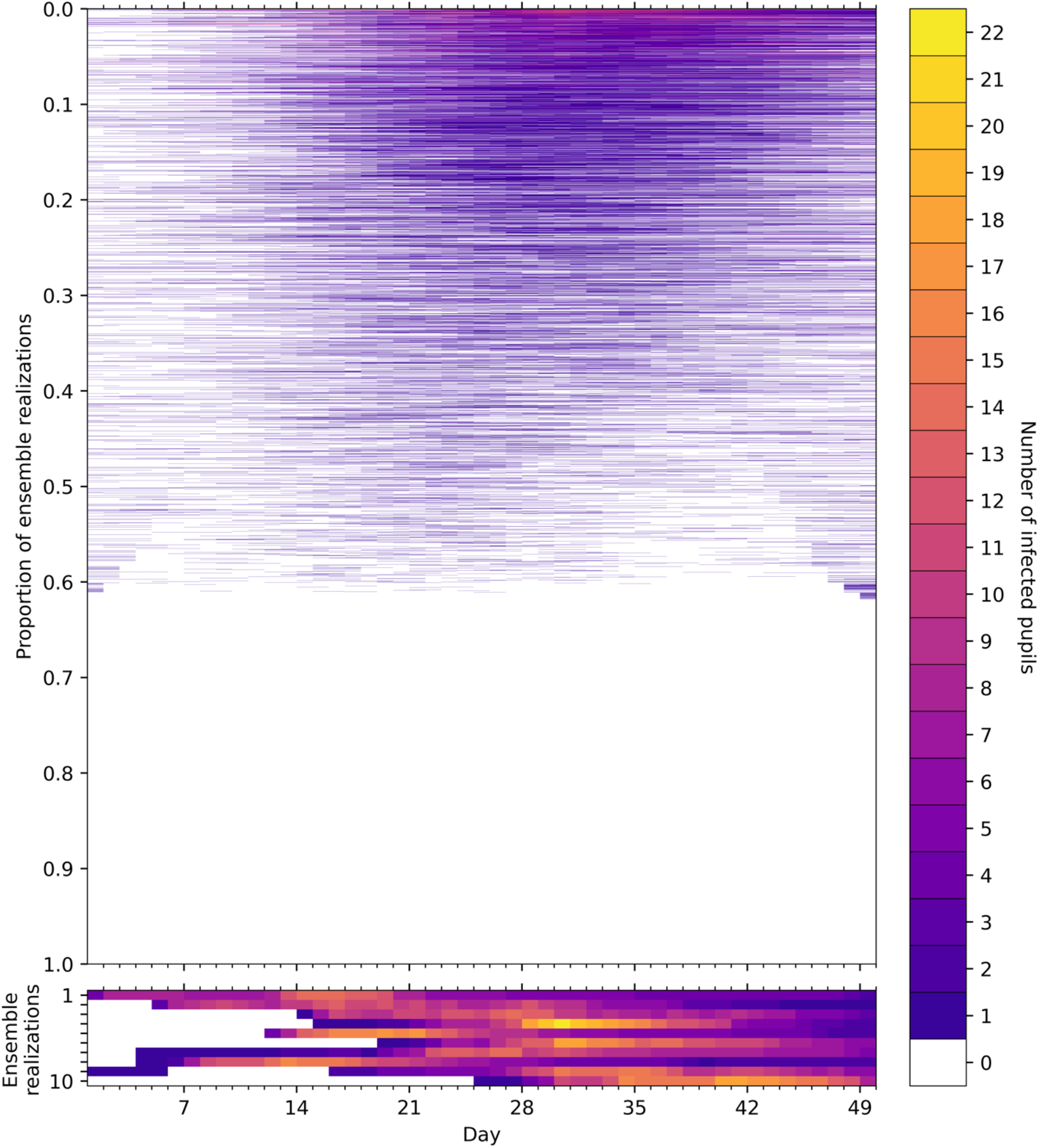
Time series of the number of infected pupils for each realization in an ensemble of 100,000 stochastic simulations of an isolated classroom in Term 1 of 2021 under the gradual-relaxation scenario. Each row of the heatmap corresponds to an ensemble member, with colours denoting the number of pupils in the classroom with Covid-19 infection on each day. The ensemble realizations have been sorted by the sum of the number of infected pupils over all days. (Upper panel) All ensemble realizations are shown. (Lower panel) The ten ensemble realizations with the most infections are shown, illustrating the dynamics of large outbreaks that occur with a frequency of 1/10,000. Colours denote the number of infected pupils in the classroom for each day of the simulation.

The gradual-relaxation scenario can also be compared to forecasts under the rapid-relaxation scenario (Figure 15) where the incidence rate is approximately three times larger at the start of Term 1. In this case, there are infected pupils in more than 89% of the ensemble realizations, and more realizations have substantial numbers of pupils simultaneously infected. This is confirmed in Figure 16 where histograms (empirical pdfs) of the total number of infected pupils in each realization are shown for both scenarios for Term 1 2021 and the equivalent simulation for Term 1 2020.

**Figure 15.**
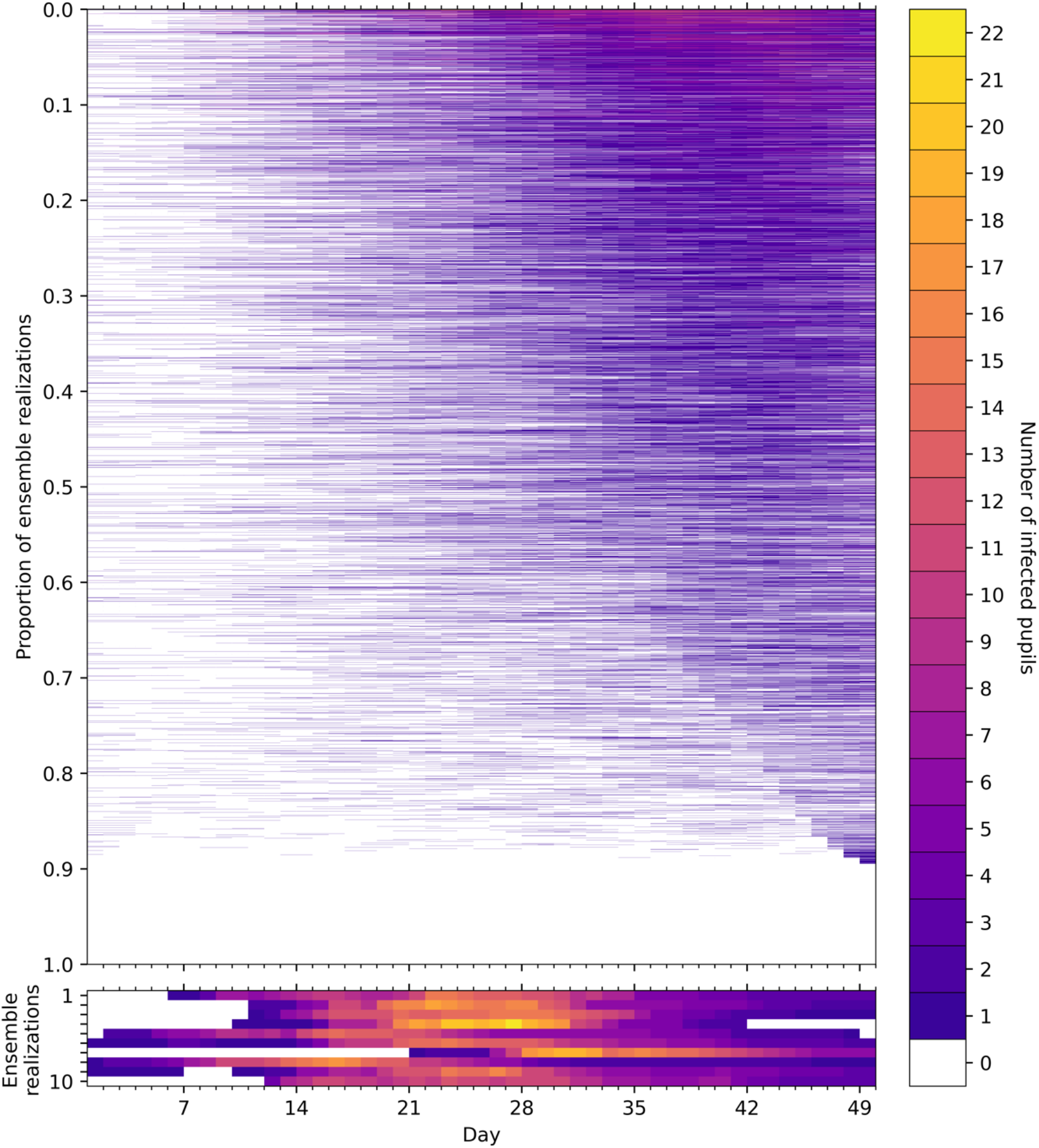
Time series of the number of infected pupils for each realization in an ensemble of 100,000 stochastic simulations of an isolated classroom in Term 1 of 2021 under the rapid-relaxation scenario. Each row of the heatmap corresponds to an ensemble member, with colours denoting the number of pupils in the classroom with Covid-19 infection on each day. The ensemble realizations have been sorted by the sum of the number of infected pupils over all days. Colours denote the number of infected pupils in the classroom for each day of the simulation. (Upper panel) All ensemble realizations are shown. (Lower panel) The ten ensemble realizations with the most infections are shown, illustrating the dynamics of large outbreaks that occur with a frequency of 1/10,000.

**Figure 16.**
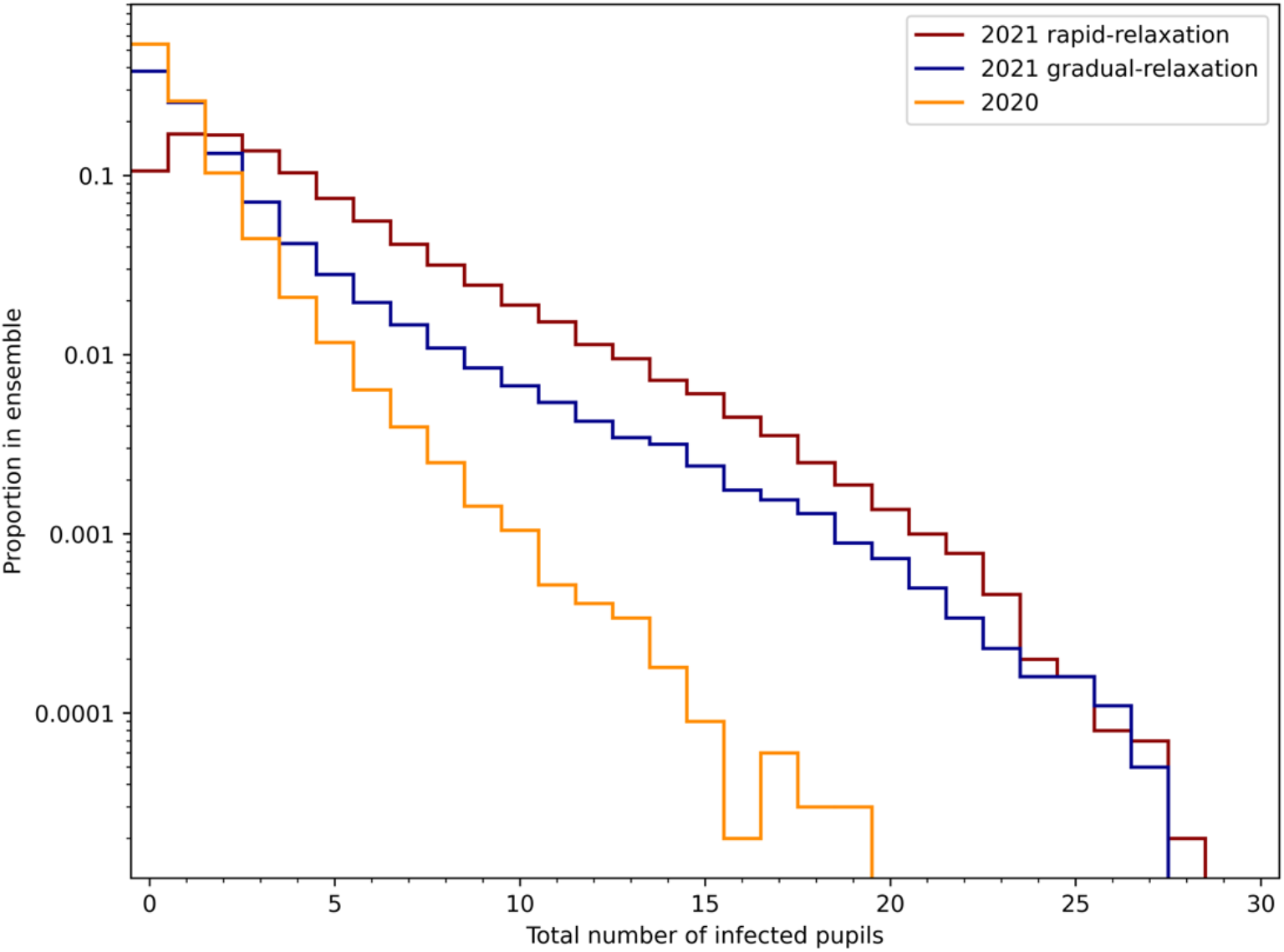
Histograms (empirical pdfs) of the total number of infected pupils in ensembles of 100,000 simulation of the rapid-relaxation and, gradual-relaxation scenarios for Term 1 in 2021 and the ‘no hard mitigations’ simulation with reduced contacts for Term 1 in 2020. Note the logarithmic scale on the proportion in the ensemble.

The results in Figure 16 show that for both scenarios for Term 1 of 2021, there is a probability of greater than 0.01 of classes having 10 or more infected pupils over the course of the term. Indeed, the number of infected pupils at the 1% probability level increases from 5 for the 2020 simulation, to 7 for the gradual-relaxation scenario for 2021, and to 13 under the rapid-relaxation scenario. The change in the upper tail values from Term 1 2020 (with no hard mitigations) to the 2021 scenarios is even more pronounced: the 99.9 percentile values on the numbers of infected pupils are 12 for 2020 and 22 for the 2021 scenarios. Thus, the simulations indicate a much greater potential for substantial outbreaks in Term 1 of 2021.

Simulations have also been performed for Term 1 of 2021 scenarios assuming the classroom contacts rates return to pre-Covid levels. To compare these simulations with those above, we consider again the collection of ten schools each with four classrooms of 30 pupils. This better illustrates the level of infection that local authorities, city boroughs or academy networks may have needed to respond to in Term 1 of 2021.

In Figure 17 we illustrate the differences that may occur in these scenarios by plotting histograms that show the probability that a given number of classes in the collection experiences infection at different levels. We find that the probabilities of classes having no infections (Figure 17a) is independent of the classroom contact rates, but strongly depends on the assumed societal contact relaxation scenario, with a rapid-relaxation to pre-Covid societal contacts substantially reducing the number of classes with no infections. Similarly, the number of classes experiencing a single infection (Figure 17b) has only slight variations with the classroom contact rates, but differ substantially under the two societal contacts forecast scenarios.

**Figure 17.**
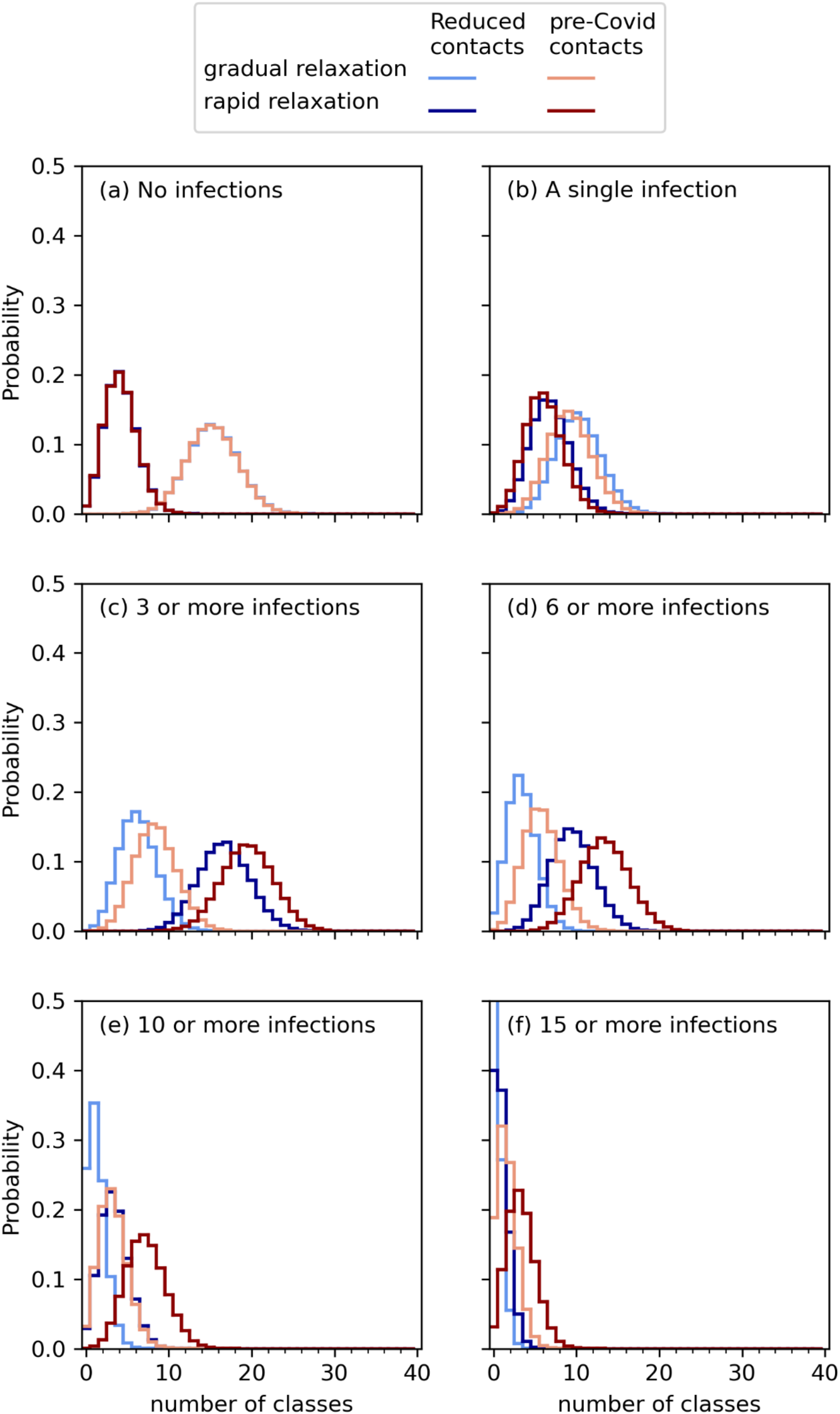
Histograms of the number of classes in a collection of ten schools each with four classes of 30 pupils that are forecast to experience SARS-CoV-2 infections in Term 1 of 2021 under different scenarios: gradual- and rapid-relaxation to pre-Covid societal contacts, and reduced or pre-Covid contact rates in the classroom. Six different infection levels are considered: (a) classes with no infections; (b) classes with a single infection; (c) classes with at least one infection; (d) classes with more than six infections; (e) classes with ten or more infections; and (f) classes with 15 or more infections. Note, in (f) the reduced contacts gradual-relaxation scenario has a probability of no classes with 15 or more infections of 0.66 and is curtailed in these plots to better illustrate the lower probability by high consequence outbreaks

The effects of in-classroom contact rates increase if we consider outbreaks. Small clusters of three or more infections (Figure 17c), which would be likely to trigger ‘extra action’ under DfE guidelines [30], are more likely for high (pre-Covid) in-class contact rates of than for reduced contacts. However, differences in the community incidence rate in the two forecast scenarios have the greater effect; three or more infections are likely in around half of the 40 classes in the collection under the rapid-relaxation scenario, but in only 5—8 classes for the gradual-relaxation scenario.

For outbreaks of more than six infections in a class (Figure 17d), our model suggests 16 classes in the collection are likely to experience such infection levels with pre-Covid classroom contact rates under the rapid-relaxation scenario, reducing to 12 classes under the gradual-relaxation scenario. Outbreaks of ten or more infections (Figure 17e) are less likely in all simulated conditions, but in the worst-case modelled (pre-Covid classroom contact rates under a rapid-relaxation of societal contacts) it remains likely that 8 classes in the collection would experience an outbreak of this size. Additionally, the number of classes with ten or more infections is similar for reduced classroom contacts in the rapid-relaxation scenario and the pre-Covid contacts in the gradual-relaxation scenario.

Considering a large outbreak of 15 or more pupils (i.e., at least half of the pupils being infected), we find that this is likely in three classes, and may occur in more than ten classes, under the worst case modelled, but becomes unlikely in the gradual-relaxation scenario. However, even in the best case modelled (reduced classroom contact rates and gradual-relaxation of societal contacts) there is an appreciable probability (approximately 0.0075) of three classes experiencing a large outbreak.

Finally, we estimate the proportion of pupils absent through Term 1 of 2021 and compare with estimates of pupil absences from reports to the Department of Education [31]. Table 5 collates the proportions of absences over six weeks in September and October 2021 as found from model ensemble simulations within the rapid- and gradual relaxation scenarios and adopting pre-Covid or reduced contact rates. We model a twice weekly rapid testing regime, both with and without confirmatory PCR testing, as well as simulations with no hard mitigations applied for comparison. We note that regular testing of primary aged children was not mandated by the UK Government during this time, although staff were required to self-administer tests twice weekly and households of pupils were recommended to carry out tests. However, an internet search has found several examples of primary schools advising regular testing of children at home, so it is likely that ad-hoc arrangements were in place in many schools.

**Table 5.** Comparison of reported pupil absences in state-funded primary schools in England with modelled pupil absences under rapid-relaxation and gradual relazation scenarios over term 1 of 2021. Model ensemble simulations with 100,000 realizations are performed adopting pre-Covid and reduced contacts distributions. Reported absences obtained from [31].

|  |  | Date | 09/09 | 16/09 | 23/09 | 30/09 | 07/10 | 14/10 |
| --- | --- | --- | --- | --- | --- | --- | --- | --- |
|  |  | Reported absences | 1.1% | 1.4% | 2.0% | 1.9% | 1.8% | 2.2% |

| Rapid relaxation scenario | pre-Covid contacts | No hard mitigations | 0.2% | 0.6% | 1.3% | 2.2% | 3.4% | 4.8% |
| --- | --- | --- | --- | --- | --- | --- | --- | --- |
|  |  | Twice weekly rapid test (no PCR) | 0.9% | 1.5% | 2.1% | 2.8% | 3.7% | 4.6% |
|  |  | Twice weekly rapid test (with PCR) | 0.2% | 0.5% | 1.1% | 1.8% | 2.6% | 3.5% |
|  | Reduced contacts | No hard mitigations | 0.2% | 0.4% | 1.0% | 1.7% | 2.6% | 3.6% |
|  |  | Twice weekly rapid test (no PCR) | 0.9% | 1.5% | 2.0% | 2.6% | 3.4% | 4.2% |
|  |  | Twice weekly rapid test (with PCR) | 0.2% | 0.45% | 0.9% | 1.6% | 2.3% | 3.1% |

| Gradual relaxation scenario | pre-Covid contacts | No hard mitigations | 0.2% | 0.6% | 1.3% | 2.2% | 2.6% | 2.3% |
| --- | --- | --- | --- | --- | --- | --- | --- | --- |
|  |  | Twice weekly rapid test (no PCR) | 0.9% | 1.5% | 2.1% | 2.6% | 2.7% | 2.3% |
|  |  | Twice weekly rapid test (with PCR) | 0.2% | 0.5% | 1.1% | 1.6% | 1.6% | 1.2% |
|  | Reduced contacts | No hard mitigations | 0.2% | 0.4% | 1.0% | 1.6% | 1.8% | 1.5% |
|  |  | Twice weekly rapid test (no PCR) | 0.9% | 1.4% | 1.9% | 2.4% | 2.5% | 2.1% |
|  |  | Twice weekly rapid test (with PCR) | 0.2% | 0.5% | 1.0% | 1.4% | 1.4% | 1.1% |

Table 5 indicates an increasing level of reported absence over the first three weeks of Term 1, almost doubling from ∼1% of pupils on 9^th^ September 2021 to 2% on 23^rd^ September 2021. Subsequently, the absence rate fluctuates at around 2% of pupils for the remaining weeks of Term 1. Our modelling results are broadly consistent with these reported values. There are differences observed in the absences between the two community prevalence scenarios, which increase over time, with approximately twice as many absences under the rapid-relaxation scenario on 14^th^ October 2021 than the gradual-relaxation scenario. As expected, reduced contact rates result in lower numbers absent.

In the first week of Term 1 (9^th^ September 2021), our modelled values are substantially lower than reported (Table 5), except in the case where twice weekly rapid testing without PCR confirmation is applied. We note here that pupils were asked to perform two self-administered lateral flow tests in the week prior to returning to school in September. This requirement was introduced after our simulations were performed, so was not included in our model, and is likely to result in model simulations under-estimating absences in the first week, but is somewhat corrected where we have modelled twice weekly rapid testing.

The modelled absences generally increase monotonically over the weeks of Term 1 under the rapid-relaxation scenario, but with gradual-relaxation the absence level shows a similar fluctuation for the final three weeks of Term 1 as seen in the reported absences (Table 5). The simulation results under the gradual-relaxation scenario with reduced contact rates and twice weekly rapid testing (with no confirmatory PCR tests) show particularly good matches to the observed rates.

## 6. Discussion

### Main results

We have developed a stochastic discrete compartmental model of SARS-CoV-2 transmission in primary school classrooms and applied this to analyse infections over Term 1 of 2020, and to forecast Term 1 of 2021. The model takes account of the known large variations in the contact patterns of individual pupils in primary schools [2]. This includes allowing for the possibility of having one or more very gregarious individuals in a mixing group, who may drive major in-class outbreaks, and allows the potential impact of such pupils to be investigated quantitatively. Our basic model is for transmission within a class, but individual schools can be modelled as collections of such classes such that the results can be applied to local authority areas and academy networks and provide estimates on a national scale.

Our analysis of model findings for Term 1 of 2020 has focussed on assessing alternative mitigation measures that have been proposed to manage infection risk in schools. While there are limited data with which to test our model, the reanalysis broadly aligns quantitatively with observations from 2020. Furthermore, in comparing mitigation measures, our model parameters are not varied, allowing relative changes to be compared and reducing the impact of uncertainties on parameters.

We find the dynamics of infection in the classroom follow the external community incidence rates through the seeding of infection. Typically, there are few instances of classroom transmission and, in the majority of ensemble realizations, there are either no infections or a single pupil is infected.

Our model results support the notion that outbreaks can be defined as infections exceeding about 5 persons in a class-sized mixing group. Below 5 cases, we find that the infections are largely driven by community prevalence, but this connection is broken for realizations numbering more than 5 infections with large outbreaks driven by classroom transmission. Our forecasts indicate that outbreaks may be substantially larger and more frequent in Term 1 of 2021 compared to Term 1 of 2020. This is due to the higher transmission rates with variant Delta SARS-CoV-2 and its more rapid onset of infectivity. In the worst-case scenario, the modelling indicates that almost all school classes will experience at least one infection case in Term 1 of 2021.

A prominent result of our modelling is the essential role of contact rates within the classroom on infection transmission. In this study, we have compared two different contact rate distributions, derived from an elicitation of teachers in May 2020 [2], using our new agent-based stochastic transmission model. The reduction of contact rates from pre-Covid levels is shown to have a comparable reduction on pupils’ infection as implementing bubble quarantine with pre-Covid contacts. The reduction of contacts also shrinks in-class secondary infections to a similar extent as the application of bubble quarantine.

Following on from these retrospective analyses, we have applied the new model to forecasting infection in schools for Term 1 2021. Here the model assumptions and parameters have been changed from the September 2020 situation to include a largely vaccinated adult population and the more virulent Delta variant of SARS-CoV-2. We have contrasted two different contact patterns, namely pre-Covid with no mitigation and then reduced contact levels, as ascribed to primary schools in Term 1 of 2020. We then contrasted two different modelling projections, concerning community prevalence and incidence rate of infection, making different assumptions about how quickly the public would alter their community contact patterns back to pre-Covid rates. In comparing these two incipient scenarios for September--October 2021, we conclude, commensurate with [6], that prevalence levels in the community will remain the dominant control on infection in primary schools.

Adopting ‘soft’ mitigation measures to reduce transmission has a modest effect compared to differences in projected community prevalence but significantly suppresses occurrence of larger outbreaks.

### Implications for managing Covid-19 in schools

Our results have implications for the management of SARS-CoV-2 infection transmission in primary schools. We find that mitigation steps that reduce contacts between pupils have a tangible benefit in disrupting transmission. Schools are already experienced in organising classrooms and other social interactions to reduce pupil-to-pupil and pupil-to-teacher contacts. Our model results suggest that, given practicalities of managing contacts within schools, continuation of these practices can serve to reduce in-school transmission by up to 30%.

Of greater significance is the comparison of different strategies to deal with infection occurrences. The exclusion of whole classrooms and bubbles has led to self-isolation of large numbers of pupils from primary schools in the recent past, with major adverse effects on education. In addition, such absences have significant and unwelcome knock-on effects on families and the economy. Our model results show that, in terms of infection numbers and transmissions, there are no tangible benefits of this policy in comparison to a policy of simply removing a pupil who has been infected from school. Thus, excluding whole classes or bubbles is hard to justify from a public health perspective and in the context if its highly disruptive effects on education and society.

Our modelling also shows that a regular rapid lateral flow testing regime has significant benefits in reducing transmission and is more effective than bubble or class exclusion. This results in much lower absence numbers, so is manifestly superior from an educational and societal perspective. Contrasts between different testing regimes with respect to frequency of testing and whether lateral flow testing is augmented by PCR tests are marginal. Our model results indeed suggest that augmented PCR tests have little benefit since a positive lateral flow result will, in most cases, simply be confirmed by a PCR test. Thus, a testing regime which meets criteria of practicality and cost should be chosen. In the context of our modelling, the findings suggest two lateral flow tests per week is a suitable regime, with little or no need for PCR tests.

Our models have limited ability to forecast, and rely on data for community incidence rates and prevalence. At the time of conducting our simulations for the return to school in autumn 20201 there were very large uncertainties in future projections of incidence rate and prevalence. Projections over long timescales (several weeks) are unreliable. In particular they are sensitive to changes in model parameters that are influenced by external forcings (e.g., weather conditions influencing the extent of indoor mixing in the summer) and changes in the characteristics of the virus that affect its tranmissivity, as is being demonstrated by the arrival of the Omicron variant. Subsequent observations are substantially different from projected community infection levels (Figure 13). Community incidence is the main driver of infection in school, seeding infection in the classroom. In our simulations, this remains the case even with a large proportion of the adult population vaccinated. We caution though that - to some extent - community prevalence control is built into our models (see next section for fuller discussion).

### Caveats, limitations and further developments

Our model makes a number of assumptions and has some limitations. We are considering only close contacts (for clarity, define here as per [2]) as an infection transmission pathway. Unavoidably, the contact pattern data we have used, derived from teacher elicitation [2], are an approximation of complex varying patterns of human interactions. Our model also assumes each classroom is isolated and independent of other classrooms, as far as transmission likelihood is concerned. Clearly there are interactions between pupils and adult staff from different classes and across age groups, but we judge this simplification is justified from evidence of limited cross age mixing, e.g. [7]. Our model also simplifies the complex social interactions by assuming that contacts are randomly distributed amoungst all members of a class, whereas there will be friendship groups resulting in variations in the connectness of the individuals with one another.

The in-classroom transmission of infection in our model is assumed to be predominantly through close contacts. This neglects the possibility of long-range airborne transmission by dispersed aerosol, which may be a contribution in situations where there are several infected people in the classroom, thereby elevating the background aerosol concentration to comparable levels to those near infectious individuals. While a model of such transmission [e.g., 37] could be incorporated in addition to the close-contact model, we note, however, that such embellishment of the model will introduce additional uncertain parameters that vary widely between schools, and is unlikely to alter the relative effectiveness of the mitigation measures.

Our model does not consider other measures to mitigate transmission (e.g., masks, cleaning and ventilation) in detail (only adopting a lumped parameter to represent these ‘soft’ mitigations). Some or all of these might be taken into account in development of a more sophisticated, more complicated model; this said, we doubt there would be much benefit for such additional complexities, given they would all introduce uncertainties and, it can be argued, would only introduce second order factors and effects.

We have not performed extensive calibration of our model, in part due to a lack of detail in published data with which to compare our model. While calibration is likely to improve our predictions of infection levels and resulting pupils absences, there is also a danger of over-fitting models as there are a number of parameters that can be tuned and data is scarse and uncertain. We have adopted published values of epidemiological parameters, where available, and have demonstrated that the model is able to reproduce observed levels of pupil absence. Even without detailed calibration, the model provides a useful tool with which to simulate potential scenarios and provide evidence to judge the effectiveness of alternative mitigation measures and the tension between controlling infection level and pupil absence.

One major issue is that the model does not explicitly distinguish between community-related infection transmission outside and inside school. Thus, to a large extent, the current model is predicated intrinsically on community prevalence being dominant. The model is, however, able to identify transmission clusters and larger outbreaks of in-school infections. We think this aspect of modelling could be usefully developed to allow it to detect differences between infection rates inside and outside school separately. With much of the adult population vaccinated, it could be that schools become important reservoirs and drivers of overall community infection.

In terms of ‘outbreaks’ of infection, at this stage we have concentrated on projections for six or more infections in a class because this breakpoint clearly emerges from the data. In this regard, in Section 4 we mention current DfE contingency operational guidance for schools [30] where two thresholds are defined for “extra action” under specific setting conditions and in relation to ‘close mixing’ groups within schools. If we take the second of these, i.e., “10% of children, pupils, students or staff who are likely to have mixed closely test positive for COVID-19 within a 10-day period”, for our hypothetical class of thirty this equates to 3 or more positives. *En passant*, in Figure 15c, we present projected number of classes - in a collection of ten schools with four classes each - that could match this concentration of infections, under two mitigation scenarios. There is, therefore, a need to focus our modelling specifically on these stated DfE criteria, and to develop such projections – and, critically, their associated uncertainties - in more detail than has been possible thus far.

Another useful potential development relates to forecasting. Forecast reliability in such models is strongly influenced by associated uncertainties. Our example of using established models of community transmission (Figure 13) indicates that their forecasting skill is limited to no more than than a few weeks. Nonetheless our modelling might usefully be combined with near real-time data assimilation of observed incidence to generate useful short-term forecasts that could be reasonably dependable, out to a few weeks ahead.

## 7. Conclusions

Substantive implications for the relative efficacies of different mitigation measures in primary schools are indicated by our agent-based contact transmission modelling results, reported above. For instance, our modelling adduces evidence that, with the emergence of the Delta variant and other changing factors, there could be a distinct contrast between the efficacies of bubbles and testing when it to comes to projected numbers of future Covid-19 infections in school and consequent absences. We contend this is a key finding for policy considerations. In addition, our results indicate potential benefits from refocusing on social distancing in schools, as well as enhanced ventilation and good hygiene.

These, and related arguments from our modelling, suggest that - in forecast mode - our stochastic contact network framework modelling should be of value to Directors of Public Health and Education Authorities charged with responsibility for prompt assessment and managing of infection transmission dynamics in school classes and in multiple schools in a given area. In terms of current or future DfE action criteria [e.g. 30, 36], our model could be tailored straightforwardly to schools’ demographics and to the relevant local community incidence level.

The numerical modelling code has been fine-tuned for rapid analytical computation and, therefore, could be used on a frequent, real-time basis under rapidly changing Covid-19 infection conditions – e.g. for the new, emerging Omicron variant; with suitable resources, daily data-adaptive updating of projections should be feasible for a group of schools in a region.

## Data Availability

Data is available on request from

## Acknowledgments

We thank Prof. Sarah Lewis (Bristol Medical School, University of Bristol) and colleagues in the “COVID-19 Mapping and Mitigation in Schools” (CoMMinS) project team for their generous advice and assistance in relation to this study.

MJW and WPA acknowledge funding from the “COVID-19 Mapping and Mitigation in Schools” (CoMMinS) project which is supported by the Medical Research Council (grant no. MR/V0285545/1) and hosted within the UK MRC Integrative Epidemiology Unit at the University of Bristol (MC_UU_00011/5). There was no direct funding for this study but the support of the Royal Society RAMP initiative for COVID-19 is acknowledged. MJW also acknowledges funding from NERC on fellowship NE/R003890/1.

## Supplementary Material

## A1. The role of airbourne transmission in close contact transmission of SARS-CoV-2

One of the emerging ideas on the spread of SARS-CoV-2 is that it occurs by airborne transmission of aerosols carrying the virus [12]. This concept has gained currency as evidence emerges for transmission in settings which require long distance transport of fine aerosols, away from an infected source, as recently summarized in [1]. The specific principles and issues related to airborne transmission in classrooms and factors related to ventilation in schools have been reviewed by Ding et al. (2022) [38]. Transmission and risk models have been developed [e.g., 37, 39, 40] which frame transmission risk in terms of long-range dispersal of aerosols within closed and open spaces. The transmission risk is a function of many parameters but, at its basic level, is controlled by the balance between recharge of a closed space with aerosols from infected persons and their removal by fresh air or ventilation. Exposure risk increases as viral concentration within the aerosols increases and is also as a function of exposure time. Aerosol size is also likely to play a role as size affects buoyancy and persistence in suspension.

In contrast, many epidemiological models, including the agent-based model in our paper, identify close contact with an infected person as the main circumstance for viral transmission to a susceptible person. This justifies social distancing as a primary approach to mitigation. Long-range and short-range exposure models to airborne aerosols are not mutually exclusive. Indeed, both kinds of process will occur concurrently in many settings [12, 38]. The question then is in what circumstances does one or the other become dominant?

Here we consider the case that short range airborne exposure is likely to be dominant in many classroom settings, supporting the development and validity of agent-based close contact models. The main concept is that various kinds of exhalation (e.g., normal breathing, talking, singing, laughing, coughing and sneezing) lead to high concentrations of virus in airborne aerosol in the immediate vicinity of an infected person [41]. Thus, the risk of being infected when close to or face-to-face with a person is greatly increased compared to the background exposure due to long range dispersal. Exhalations from an infected person create virus-laden jet plumes that involve turbulent entrainment of air, so the aerosol becomes diluted as the plume disperses. Relevant research has been recently reviewed in Burridge et al. [42]. Abkarian et al. (2020) [41] and Chen et al. (2020) [43] provide quantification of typical dilution rates as a function of distance in exhalations. As examples, the concentration of aerosols in a turbulent jet produced by speech reduces to 6% at 1m and 3% at 2m from its initial value [41]. Concentrations of fine aerosols significantly above background are calculated at up to 0.3 m for speech (counting) and up to 1 m for coughing [43]. Substantial distances, greater than typical classroom sizes, are required for concentrations in the respiratory jet to reach background levels [37]. Because proximity (∼ 1 m) to an infected person is now a recognised major risk factor, these results, and the illustration below, support our model choice of close contact as the predominant mechanism in a school classroom.

In order to appraise the relative risk of infection to pupils in the room, away from close contacts, we can compare the background concentration of infected aerosol with the local concentration close to an infected person. Let us take 6 litres per minute of air inhaled as typical of both an adult female and a primary age child (with adult males 50% higher) [44] and assume the concentration of virus-bearing aerosol of an infected child is some unknown value X. Taking a classroom of 8 × 6 m as the minimum in UK Primary schools with height 3 m we have a volume of 144,000 litres. We now need a suitable time scale and choose 3 hours, noting that this is the duration of a school half day and that the half-life of the virus is 1.1 hours [45]. Thus, the room cannot have a concentration more than 0.0075X. However, the concentration will be very much less if there is ventilation. According to [38] a classroom of 30 children should have a room ventilation rate of at least 120 litres per minute, meaning that it would take only 20 minutes to replace the air in a classroom. Here the background concentration of infected aerosol would be approximately 10^−3^ X. Thus, concentrations of virus in aerosols, and therefore risk of infection, can be tens to hundreds of times greater than background within a metre of an infected person. We conclude that, in most circumstances, close contacts will dominate transmission in a primary school classroom, and so justifies our choice of model. We acknowledge that risk from long range airborne transmission could become much more significant in situations of poorly ventilated classrooms and with multiple infected pupils during an outbreak.

## A2. Details of the agent-based classroom transmission model

Agent-based models (ABMs) are a convenient, powerful and flexible approach to simulation of infectious diseases [46]. Of particular relevance to our application, ABM are useful in small populations where there is heteorogeneity in population characteristics. Furthermore, ABMs can be easily combined with network models, including where the network evolves over time [e.g. 47].

We develop a simple, bespoke ABM to model SARS-CoV-2 transmission in contacts between people in a school classroom. Our agents are individuals in the classroom, which include pupils and a teacher, and could be extended to include teaching assistants and other school staff members. Each agent has their own characteristics, drawn from specified probability distributions.

In our current application, we consider only pupils and teachers in the classroom, and therefore there are two classes of agents. The characteristics of the two classes are similar, except that teachers can be permanent or temporary, but the parameters and distributions vary between pupils and teachers, reflecting differences between adults and children.

Steps in modelling an isolated classroom

1. Initialize classroom
  i. Specify size of class, *N*_*pupils*_
  ii. Initialize *N*_*pupils*_ agents in pupil class using SEUQR Initialization routine
  iii. Initialize single agent in teacher class using SEUQR Initialization routine
  iv. If Teacher is in Quarantine on initialization then replace teacher with Temporary Teacher
2. For each day of simulation, do
  i. Update community infections in classroom: for each Susceptible agent in the classroom, do:
    a. set community transmission probability from time series of community incidence rate at current time step;
    b. if agent is vaccinated then reduce transmission probability using Vaccine susceptibility factor;
    c. perform Bernoulli trail using transmission probability – agent is infected if trail is successful;
    d. if agent is infected, start infection time counters.
  ii. If rapid testing is implemented and current day is a test day then, for each agent, do:
    a. if agent is not Unwell and agent has not tested positive in PCR test, then perform a rapid lateral flow test as a Bernoulli trial with probability of success given by:
    - rapid test sensitivity if agent is infected;
    - 1 – rapid test specificity if agent is not infected. Rapid test is positive if Bernoulli trial is ‘success’.
  iii. If Bernoulli trial is success, then:
    - if Bubble Quarantine is applied on positive rapid test, then more all agents to Quarantine, otherwise move agent to Quarantine;
    - start quarantine counter
    - if agent is Teacher, replace with temporary teacher;
    - if agent is Temporary teacher, replace with a new temporary teacher.
  iv. If confirmatory PCR testing is implemented then, for each agent, do:
    a. if agent has retuned a positive rapid test and current time step is two days after day of agents’ latest positive rapid test, then perform a PCR test as a Bernoulli trial with probability of success given by:
      - PCR test sensitivity if agent is infected;
      - 1 – PCR test specificity if agent is not infected. PCR test is positive if agent is ‘success’.
    b. If Bernouilli trial is ‘success’, then:
      - if Bubble Quaratine is applied on positive PCR test, then move all agents to Quarantine,
    c. If Bernoulli trial is ‘fail’, then return agent to classroom and reset Quarantine counters. If agent is permanent teacher, remove temporary teacher.
    d. If class is not currently in Bubble Quarantine and current time step is a school day, then model contact transmissions in the classroom. If at least one agent in the classroom is currently infected, then for each agent current not in Quarantine, do:
      a. build contact network graph using daily contacts of each agent as the degrees of the graph nodes;
      b. loop through the edges of the contact network;
      c. if neither of the agents is infected, then skip edge;
      d. if both agents are infected, then skip edge;
      e. if one agent is infected and the other is Susceptible, then:
      - get probability of transmission from infected agent, evaluating from their relative transmissibility function at the current time step, multiplying by the agents’ transmission probability and the agents’ Soft mitigation factor;
      - if Susceptible agent is vaccinated, then reduce probability of transmission by Vaccine susceptibility factor;
      - perform Bernoulli trial for transmission in contact using the probability of transmission in this contact, with ‘success’ corresponding to infection transmission to Susceptible agent;
      - if Bernoulli trial is a ‘success’ then move Susceptible agent to Exposed and update counters.
  v. Update SEUQR compartments using SEUQR update routine.
  vi. Update Quaratine counters.
  vii. Update current day.

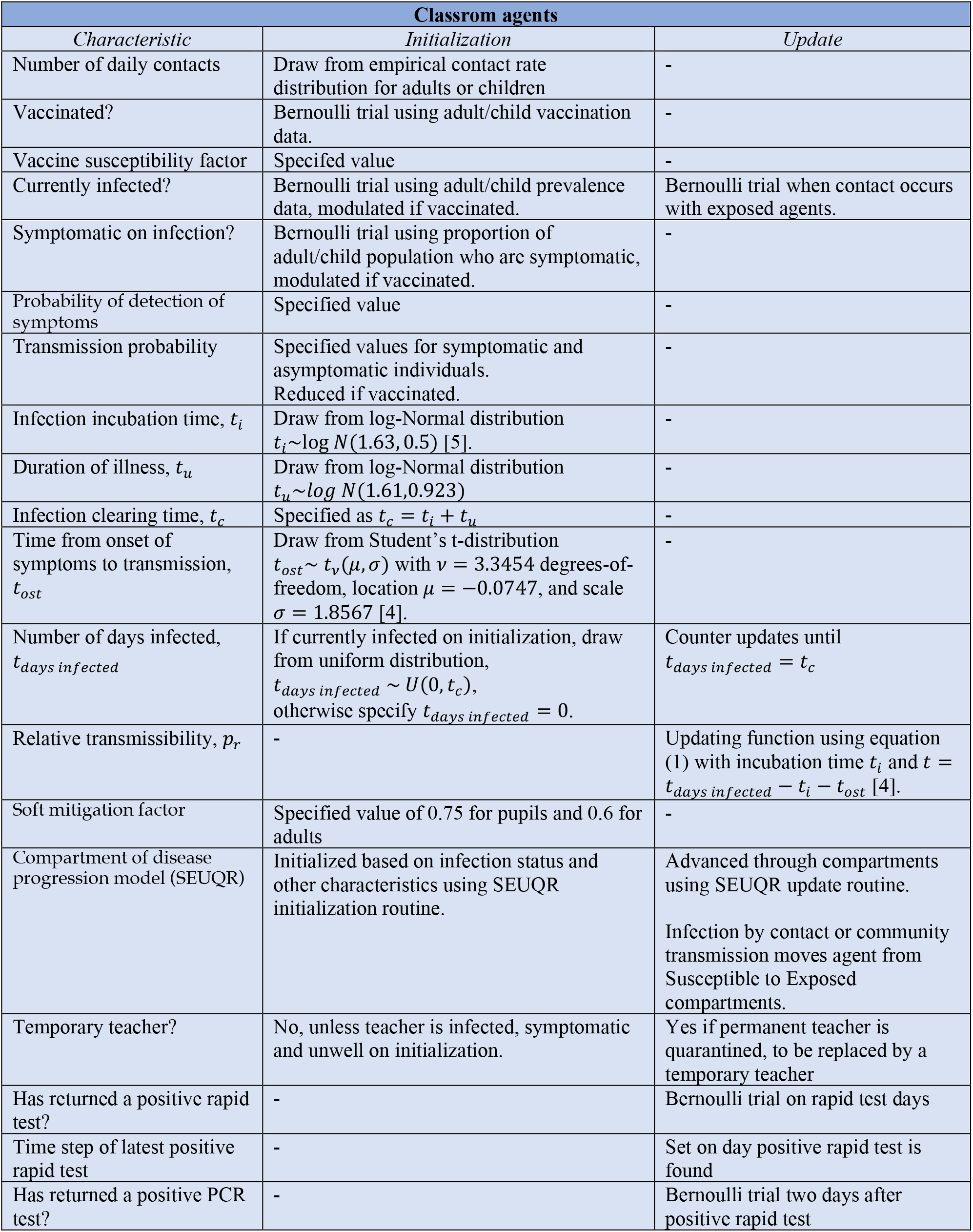

| Classrom agents |  |  |
| --- | --- | --- |
| Characteristic | Initialization | Update |
| Number of daily contacts | Draw from empirical contact rate distribution for adults or children | - |
| Vaccinated? | Bernoulli trial using adult/child vaccination data. | - |
| Vaccine susceptibility factor | Specified value | - |
| Currently infected? | Bernoulli trial using adult/child prevalence data, modulated if vaccinated. | Bernoulli trial when contact occurs with exposed agents. |
| Symptomatic on infection? | Bernoulli trial using proportion of adult/child population who are symptomatic, modulated if vaccinated. | - |
| Probability of detection of symptoms | Specified value | - |
| Transmission probability | Specified values for symptomatic and asymptomatic individuals.<br>Reduced if vaccinated. | - |
| Infection incubation time, $t_i$ | Draw from log-Normal distribution $t_i \sim \log N(1.63, 0.5)$ [5]. | - |
| Duration of illness, $t_u$ | Draw from log-Normal distribution $t_u \sim \log N(1.61, 0.923)$ | - |
| Infection clearing time, $t_c$ | Specified as $t_c = t_i + t_u$ | - |
| Time from onset of symptoms to transmission, $t_{ost}$ | Draw from Student's t-distribution $t_{ost} \sim t_v(\mu, \sigma)$ with $v = 3.3454$ degrees-of-freedom, location $\mu = -0.0747$, and scale $\sigma = 1.8567$ [4]. | - |
| Number of days infected, $t_{days\ infected}$ | If currently infected on initialization, draw from uniform distribution, $t_{days\ infected} \sim U(0, t_c)$, otherwise specify $t_{days\ infected} = 0$. | Counter updates until $t_{days\ infected} = t_c$ |
| Relative transmissibility, $p_r$ | - | Updating function using equation (1) with incubation time $t_i$ and $t = t_{days\ infected} - t_i - t_{ost}$ [4]. |
| Soft mitigation factor | Specified value of 0.75 for pupils and 0.6 for adults | - |
| Compartment of disease progression model (SEUQR) | Initialized based on infection status and other characteristics using SEUQR initialization routine. | Advanced through compartments using SEUQR update routine.<br>Infection by contact or community transmission moves agent from Susceptible to Exposed compartments. |
| Temporary teacher? | No, unless teacher is infected, symptomatic and unwell on initialization. | Yes if permanent teacher is quarantined, to be replaced by a temporary teacher |
| Has returned a positive rapid test? | - | Bernoulli trial on rapid test days |
| Time step of latest positive rapid test | - | Set on day positive rapid test is found |
| Has returned a positive PCR test? | - | Bernoulli trial two days after positive rapid test |

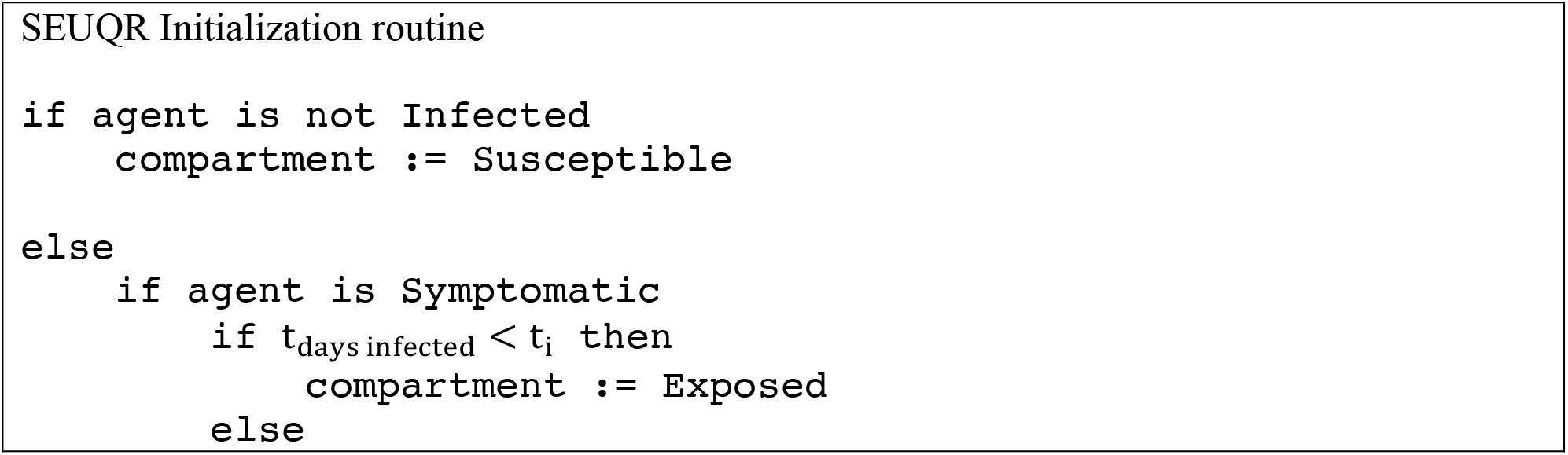
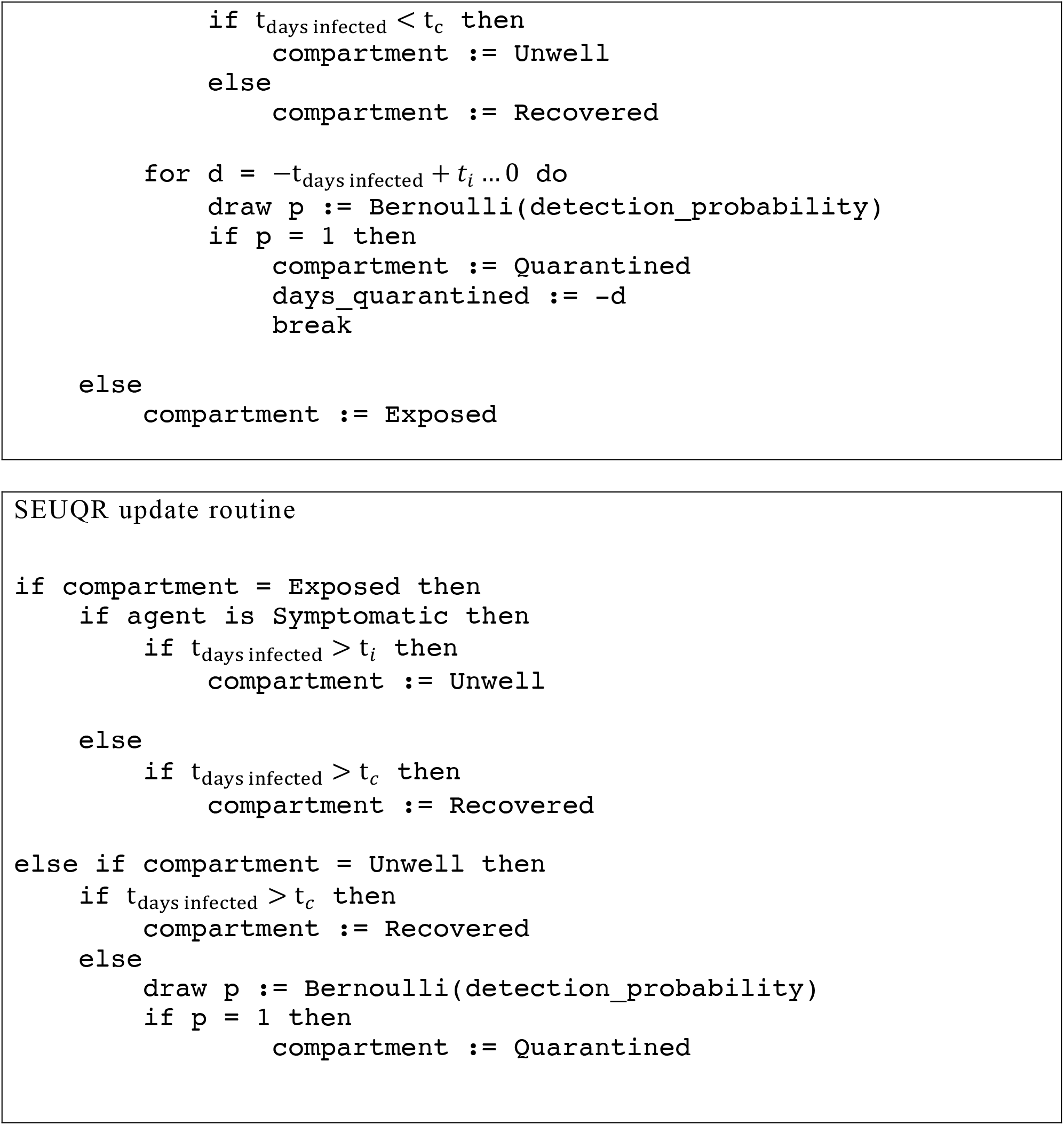

### A3. Linking prevalence to incidence rate

The roadmap projections [19-21] that we use to forecast potential future infections in schools contain trajectories for the incidence rates through Term 1 of 2021, but our model also requires time series for the community prevalence. Here we adopt a simple relationship between incidence rate and prevalence, as described here.

Our relationship is based on the SIR compartmental epidemiological model [48] that relates the numbers of susceptible (S), infected (I) and recovered (R) people in the population. New infections occur at a rate depending on the number of contacts between susceptible and infected people, with a parameter *β* (given by the product of the contact rate and probability of transmission in a contact).

Infected people recover (or are otherwise removed) with probability of recovery in time interval *dt* given by *γdt* (which is equivalent to assuming duration of infection is drawn from an exponential distribution, with mean duration *D* = 1/*γ*).

The movement of people between compartments is encoded in a system of differential equations [48], given by

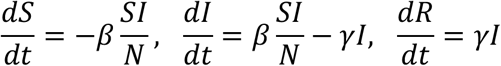

where *N* = *S* + *I* + *R* is the total population (here assumed constant). The second of these equations is pertinent to understanding the link between incidence rate and prevalence. Indeed, this equation can be written in the form

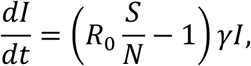

where *R*_0_ is the basis reproduction number, and where *I* is the community prevalence and *dI*/*dt* is the incidence rate of new infections in the population. For SARS-CoV-2, we simplify this expression further by noting that the data suggests that the infected population at any time is a relatively small proportion of the total population in the UK, and that recovery does not appear to prevent reinfection. Therefore we can make the approximation that *S*/*N* ≈ 1, so that

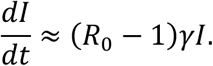

If the incidence rate *dI*/*dt* is known, and there are estimates of *R*_0_ and *γ* = 1/*D* then we can directly estimate the prevalence as

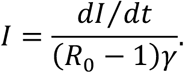

This relationship is a simple rescaling of incidence rate to obtain prevalence and suggests that high incidence rates occur for large values of the reproduction number *R*_0_ for a given prevalence (as expected), and that high prevalence occurs at a given incidence rate if the duration of infection is increased (as expected).

In using the expression derived above, we assume that *R*_0_ and *γ* are constants. This means that contact rates are unchanged, so in analysis of past data we must take periods where social contact restrictions are unchanged. Additionally, the probability of transmission in a contact must be constant, so it is unlikely to apply in periods where there are multiple variants of SARS-CoV-2 competing within a population.

We test this relationship using data from the Office for National Statistics Coronavirus Infection Survey [29], adopting daily estimates of the infection prevalence and incidence rate. The methods used for the estimate in this dataset have changed over the course of the epidemic, and therefore the interpretation of the parameter *γ* may change. In 2020, the incidence and prevalence estimates were mainly derived from random PCR testing surveys, and therefore the duration of infection *D* = 1/*γ* is likely to refer to the duration for which PCR testing detects virus in a sample. This may be substantially longer than the duration of symptoms or the duration of infectivity. Figure 18 shows the relationship between prevalence and incidence over the period 1 September to 22 November 2020. In the early part of this period (up to early-October 2020), incidence and prevalence increased and a clear linear relationship is observed. In this time, some social mixing restrictions were introduced, but do not substantially alter the trend. However, from early October 2020, there was a marked increase in the prevalence with only small changes in the incidence rate which continued until a second national lockdown was announced on 31 October 2020 and enacted on 5 November 2020. While this increased prevalence could be explained by reductions in *R*_0_ while the incidence rate remained steady, this seems unlikely, and may be better explained by a breakdown of the simple modelling assumptions during a period of change in social behaviour, e.g., a lag between prevalence and incidence that is not included in the simple SIR model. Additionally, the Alpha variant (B.1.1.7) was known to be circulating in the UK in this time.

**Figure 18.**
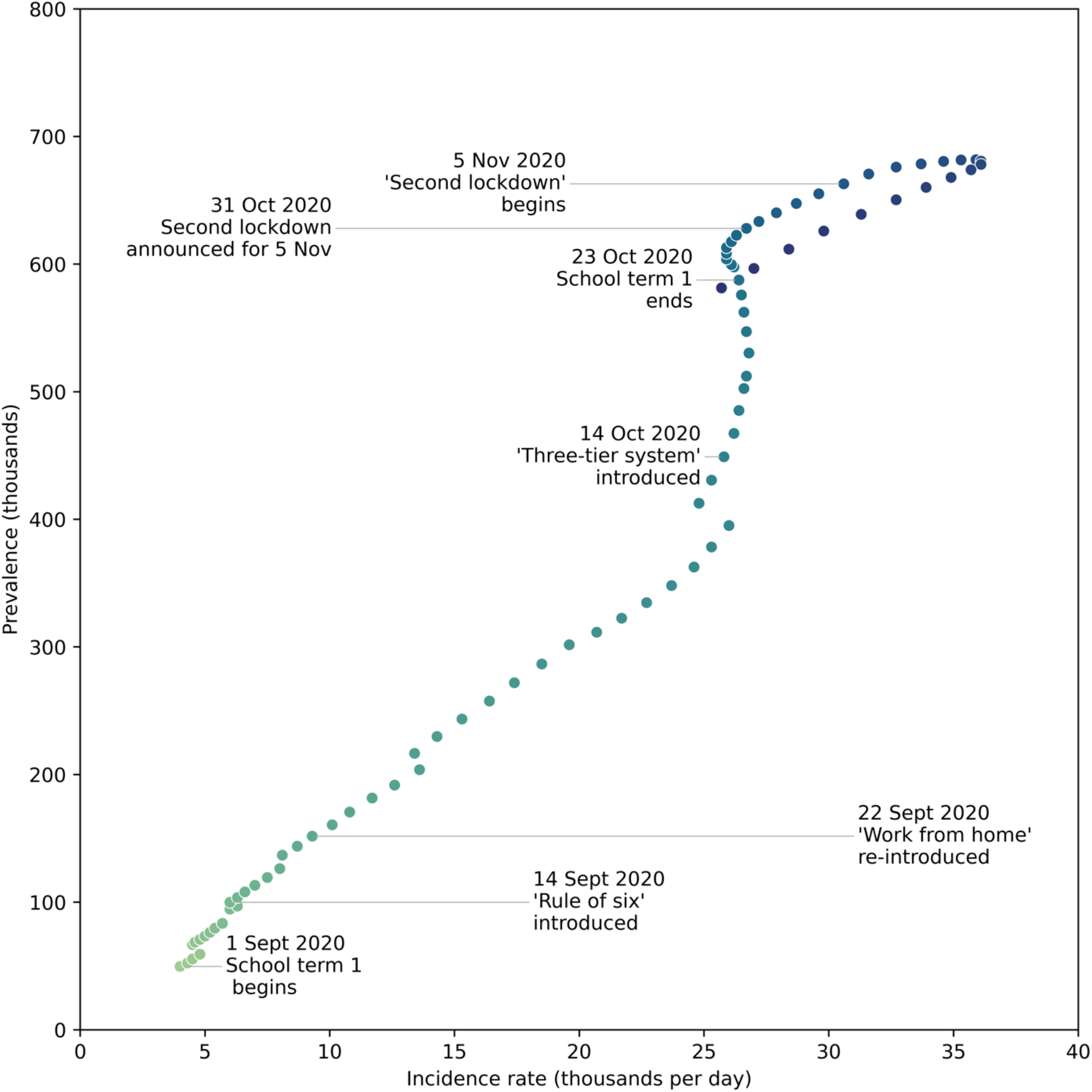
Estimates of the UK SARS-CoV-2 prevalence as a function of incidence rate for 1 September 2020 to 22 November 2020 (date from [29]). The colour of points indicates progression through time. Key dates for changes in social contact rates are indicated. Note, the Alpha variant (B.1.1.7) was first detected in a PCR sample taken in mid-September 2020 and may have been spreading rapidly in this time.

Despite these limitations, the rescaling relationship is a pragmatic and practical tool for estimating prevalence from incidence during periods where social contacts are not changing substantially. We therefore adopt this for estimating prevalence from projections of incidence rate for our forecasts in Term 1 of 2021. However, this requires estimation of the *R*_0_ and *γ* parameters. While a Bayesian approach would be valuable to quantify uncertainties, for our purposes we are content using a simple manual fitting procedure, using estimates of *R*_0_ for the dominant Delta variant (given in the range 5— 7 in [20]) and noting that increases in this value can be compensated by decreases in *γ*. This approach produces Figure 13 from ONS data for June—August 2021 and the incidence rate forecasts of [19].

## Notes

### Competing Interest Statement

The authors have declared no competing interest.

### Author Declarations

University of Bristol

### Summary of Updates

New appendix discussing the role of airbourne transmission in a classroom context. Amended figure 3.

